# Development and validation of early warning scores predicting 24-hour mortality addressing intercurrent medical interventions: a multi-centre retrospective cohort study of 2·08 million hospital encounters

**DOI:** 10.1101/2025.10.12.25337794

**Authors:** Alexandros Katsiferis, Neil Scheidwasser, Tri-Long Nguyen, Theis Lange, Mark P Khurana, Pernille B Nielsen, Kasper Karmark Iversen, Christian S Meyhoff, Eske Kvanner Aasvang, Jesper Mølgaard, Adrian G Zucco, Tibor V Varga, Samir Bhatt

**Author notes:** **Corresponding Author Details:** Alexandros Katsiferis, PhD, Section of Health Data Science & AI, Department of Public Health, University of Copenhagen, Copenhagen, Denmark Address: Bartholinsgade 6Q, DK-1356 Copenhagen K Denmark. **Author Details: Neil Scheidwasser**, Work-Address: Bartholinsgade 6Q, DK-1356, Copenhagen K, Denmark **Tri-Long Nguyen**, Work-Address: Bartholinsgade 6Q, DK-1356, Copenhagen K, Denmark **Professor Theis Lange**, Work-Address: Øster Farimagsgade, 1353 Copenhagen K, Denmark **Mark P Khurana**, Work-Address: Bartholinsgade 6Q, 1356 Copenhagen, Denmark **Pernille B Nielsen**, Work-Address: Blegdamsvej 9, 2100 Copenhagen, Denmark **Professor Kasper Karmark Iversen**, Work-Address: Borgmester Ib Juulsvej 1, 2730 Herlev, Denmark **Professor Christian S Meyhoff**, Work-Address: Bispebjerg Bakke 23, DK-2400 Copenhagen, Denmark **Professor Eske Kvanner Aasvang**, Work-Address: Blegdamsvej 9, 2100 Copenhagen, Denmark **Jesper Mølgaard**, Work-Address: Blegdamsvej 9, 2100 Copenhagen, Denmark **Adrian G Zucco**, Work-Address: Bartholinsgade 6Q, DK-1356, Copenhagen K, Denmark **Tibor V Varga**, Work-Address: Bartholinsgade 2, building 22, DK-1356, Copenhagen K, Denmark **Professor Samir Bhatt**, Work-Address: Bartholinsgade 6Q, DK-1356, Copenhagen K, Denmark.

## Abstract

**Background:** The National Early Warning Score (NEWS) is widely used to predict patient deterioration, but retrospective validations face intervention bias and emphasise discrimination over clinical utility. We aimed to compare NEWS’s clinical net benefit against simplified scoring rules and machine learning, to determine whether simpler approaches suffice or if complex models offer meaningful advantages, and to evaluate performance heterogeneity across patient subgroups.

**Methods:** We included fifteen Danish hospitals with over 2·08 million hospital encounters representing 825200 unique patients over five years (2018 to 2023). We compared NEWS against both simpler and more complex approaches for predicting 24h mortality: Simplified NEWS (NEWS without blood pressure and temperature), DEWS (Simplified NEWS with age and sex), and a model based on eXtreme Gradient Boosting (XGB-EWS) incorporating vital signs, demographics, laboratory markers, plus medical history embeddings extracted using sentence transformers. We used propensity score weighting to mitigate intervention bias and evaluated performance using net benefit, Area Under the Receiver Operating Characteristic Curve (AUC), and calibration.

**Findings:** Decision curve analysis in the overall population showed maximum net benefit differences of 1·9 additional correct mortality identifications per 10000 patients between XGB-EWS and NEWS, and 1·5 per 10000 between NEWS and Simplified NEWS. However, stratified analyses revealed substantial heterogeneity: in the high-risk group (initial NEWS ≥7), XGB-EWS provided a maximum gain of 78·1 correct identifications per 10000 patients compared to NEWS, while Simplified NEWS resulted in a maximum performance loss of 60·2 per 10000. Conversely, in low-risk groups, differences were minimal.

**Interpretation:** Model complexity produced limited average clinical utility gains, concentrated among patients initially classified as high risk by NEWS. These findings support prospective trials of risk-stratified monitoring based on the first NEWS assessment, testing simplified approaches for lower-risk patients while retaining full NEWS or electronic health-record models for higher-risk patients.

**Funding:** Novo Nordisk Foundation

**Research in context:** 

**Evidence before this study:** Early warning score systems have evolved from single vital sign monitoring to standardized multivariable scores such as the National Early Warning Score (NEWS), to even more sophisticated machine learning and deep learning frameworks utilizing electronic health records data. All these early warning scores are primarily designed to guide clinical decision-making by helping identify patients at risk of clinical deterioration within hospitals. Despite these advances, validation studies predominantly focus on statistical metrics measuring discrimination performance rather than meaningful clinical utility. On May 23, 2026 we searched PubMed for studies without language limitations published from May 23, 2016, to May 23, 2026, using terms including “early warning score”, “NEWS”, “NEWS2”, “machine learning”, “artificial intelligence”, “decision curve analysis”, “net benefit”, “clinical utility”, and “24-hour mortality.” While most of the published models, including the more sophisticated machine learning ones, have demonstrated better discrimination compared to traditional early warning scores, we found only one study that combined early warning score validation with clinical utility analysis for short-term clinical deterioration. Additionally, no studies have evaluated clinical utility across multiple patient subgroups while using causal inference approaches to address intervention bias in a large-scale healthcare system validation.

**Added value of this study:** The current work is a large-scale multi-center early warning score validation study, encompassing 2·08 million hospital encounters across 15 Danish hospitals and nine distinct clinical specialties over five years. We used the *predictimand* framework with causal inference methods to address intervention bias, a critical methodological advance in early warning score assessment. Unlike previous research focused primarily on discrimination metrics, we evaluated clinical utility using decision curve analysis across multiple patient subgroups, revealing heterogeneity in net benefit differences. In the overall population, sophisticated machine learning approaches provided marginal improvements (up to 1·9 per 10000 patients) despite superior discrimination, and simplified NEWS (without blood pressure and temperature) achieved comparable net benefit to full NEWS. However, in high-risk subgroups, particularly patients with high initial NEWS scores, differences increased substantially (up to 78·1 per 10000 for machine learning versus NEWS and 60·2 per 10000 for NEWS versus simplified NEWS), indicating that implementation strategies should be tailored to clinical context rather than uniformly applied.

**Implications of all the available evidence:** We demonstrated that differences in mortality prediction varied substantially by baseline risk: less than 2 per 10000 patients in the overall population, but up to 78 per 10000 in high-risk subgroups such as patients with initial high scores of NEWS. These findings challenge uniform implementation approaches: for general ward screening where most patients are at low baseline risk, simplified scoring achieves comparable utility while potentially enabling workflow optimization. However, for high-risk populations where baseline mortality is elevated, more complex models provide larger incremental gains that may justify additional implementation complexity. Future research should conduct prospective cluster-randomized trials evaluating context-specific implementation strategies, comparing simplified versus complex approaches across different care settings and baseline risk profiles.

## Introduction

Early warning systems are important components of modern healthcare services, providing standardized approaches to identify patients at risk of clinical deterioration.^1^ Following its development by the Royal College of Physicians in 2012, the National Early Warning Score (NEWS) was implemented throughout National Health Service hospitals and has been adopted or validated in healthcare systems worldwide, including its implementation in Danish hospitals.^1–8^ The score ranges from 0 to 20 and consists of seven vital sign parameters: respiratory rate, oxygen saturation, supplemental oxygen requirement, systolic blood pressure, pulse rate, level of consciousness, and temperature. Each parameter is assigned points based on deviation from normal values, with higher scores indicating greater risk of adverse outcomes within 24 hours. NEWS operates clinically through score-based escalation protocols: scores of 5-6 trigger urgent responses while scores ≥7 mandate emergency assessment.^8^ These protocols are predicated on the assumption that higher scores identify patients at meaningfully elevated risk of clinical deterioration. In parallel with scoring systems, such as NEWS, there has been interest in developing statistical and machine learning models that leverage broader electronic health records for predicting clinical deterioration.^1,4,7,9,10^ The latter typically incorporate laboratory results, medication and comorbidity histories, and procedural data to enhance prognostication beyond that provided by vital signs alone.

Retrospective studies of Early Warning Scores (EWSs) face methodological challenges that could undermine their clinical validity. One limitation is intervention bias: when effective early warning systems prompt interventions that rescue many high-risk patients, retrospective analysis can become problematic.^11–13^ In practice, if high-risk patients receive effective care and survive, then their observed outcome (higher survival, that is, lower mortality) deviates from the initial baseline prediction, due to the rescuing intervention in itself. Thus, retrospective analysis may create misleading associations, potentially making high scores appear to overestimate mortality, with the latter being more prominent as healthcare systems become more responsive to EWSs.^14^ This represents a core misalignment between how EWSs function clinically (as pre-intervention risk stratification tools) and how they are evaluated retrospectively (based on post-intervention outcomes). The pertinent clinical question – “What would the patient’s risk trajectory be without intervening on them?” – cannot be answered using approaches that measure observed outcomes after treatments have been administered. Beyond intervention bias, the literature has methodological gaps in performance measurement. While recent guidelines, such as TRIPOD+AI, now emphasize the importance of clinical utility metrics such as net benefit, the vast majority of validation research has focused on discrimination measures such as Area Under the Receiver Operator Characteristic Curve (hereinafter referred to as AUC) that are not designed to be interpretable in clinical settings, cannot capture real-world clinical value or misclassification costs.^1,7,15–19^ Moreover, existing subgroup evaluations remain limited to age- and disease-specific strata using discrimination metrics rather than clinical utility measures,^4,20–22^, while evidence of practical implementation value, including cost-effectiveness and feasibility, remains scarce.^23^

To inform quality improvement of early warning monitoring across the participating Danish hospitals, we conducted a large-scale multi-centre evaluation of NEWS as currently implemented in routine clinical care, comprising 2·08 million hospital encounters across 15 Danish hospitals. We employed causal prediction approaches to mitigate intervention bias and estimate EWS performance in the absence of clinical interventions. Our primary aim was to evaluate whether NEWS (standard of care across the study’s hospital network) and alternative approaches effectively predict 24-hour mortality risk, the ultimate consequence of clinical deterioration that current score-based escalation protocols are designed to detect. We thereby provide a quantitative basis that could inform potential modifications to current monitoring practice. We compared NEWS against developed simplified variants and machine learning approaches, using clinical utility metrics to assess whether score thresholds identify clinically meaningful differences in mortality risk for individuals with hospital encounters. Secondary aims included evaluating whether predictive performance varied across different patient subgroups, to support context-specific implementation decisions within the participating hospital network.

## Methods

### Study Design & Data

We conducted a multi-center retrospective cohort study, using data from 15 hospitals (10 hospital networks) across the Capital and Zealand region of Denmark, between October 16, 2018, and October 16, 2023, including a diverse range of clinical settings, specialties, and patient populations. These two regions cover approximately 46% of the Danish population (2·76 million people), spanning urban and less densely populated areas. Demographic information (date of birth and sex) and clinical data were extracted from the hospital’s electronic health records, with patient-level linkage enabled by the unique Danish personal identification number. All centers routinely collected NEWS data as part of standard clinical care during the study period. All patients were followed until death or completion of the 24-hour observation period after the initial NEWS score was recorded, whichever occurred first, with no patients lost to follow-up. We define our index time (time zero of prediction) as the time of the first NEWS measurement, collected as a specific timestamp of day and time to the minute. The study adheres to the TRIPOD+AI guidelines.

### Participants

We included all adult individuals (≥18 years) with a hospital encounter at the participating hospitals during the study period who had at least one NEWS assessment recorded. We excluded hospital stays of 15 minutes or less to ensure an observation window consistent with the clinical purpose of early warning monitoring and to minimize the inclusion of potential administrative registrations. For individuals with multiple measurements taken during a single encounter, we used the first one to establish a baseline risk before clinical interventions. Individuals could contribute multiple separate hospital encounters to the study, with each encounter analyzed as a distinct clinical prediction instance.

### Outcome

The outcome used was all-cause mortality within 24 hours (based on timestamps of date and time to the minute) of the initial NEWS recording. Time-of-death timestamps were obtained from Sundhedsplatformen, the regional electronic health record system from which all data in this study were drawn and originate from the electronic death certificate (Appendix p 1 for timestamp characteristics). For the primary analysis, we refrained from using a composite including Intensive Care Unit (ICU) admission, as the latter is influenced by external factors such as bed availability, local policies, and physician subjectivity. Instead, ICU was treated as an intercurrent intervention. We opted for 24 hours prediction as it represents the timeframe for which NEWS was originally designed, is the most frequently evaluated period in EWS validation studies and is clinically relevant for the intended use of these systems, which aim to identify patients at imminent risk of deterioration.^1,14^

### Predictors

We extracted vital sign parameters routinely collected during clinical care: respiratory rate, oxygen saturation, supplemental oxygen, systolic and diastolic blood pressure, pulse rate, level of consciousness, and temperature. Laboratory parameters included hematological markers (hemoglobin, leukocytes, and platelets), biochemical indicators (creatinine, alanine aminotransferase, lactate dehydrogenase, albumin, and C-reactive protein), and specialized tests (arterial and venous lactate and troponin T). For patients with multiple measurements of the same marker, we calculated the median across all measurements of that specific test from the patient’s medical history prior to the time of the initial NEWS recording. We used the Danish Healthcare Classification System (*Sundhedsvæsenets Klassifikationssystem* (SKS) codes) to acquire data on previous diagnoses and medical procedures from prior hospital encounters, also including ICU or respiratory support episodes. Both laboratory data and clinical history variables were available from 2018 onwards due to data system implementation. Additional predictors included demographic variables (age, sex), hospital of encounter, number of previous hospital encounters, and temporal variables related to the NEWS recording (day of week, time of day, month). We further captured SKS codes registered within 24 hours after the initial NEWS measurement to define exposure to major interventions for subsequent propensity score estimation analyses. A comprehensive description of the missing data and their handling is provided in the Appendix (pp 2, 8).

### Analytical Methods

We processed the clinical history text in Danish from previous diagnoses and medical procedures using text embeddings, which are high-dimensional numerical representations of textual data. Specifically, we used the *“minishlab/potion-multilingual-128M”* model, a static embedding sentence transformer distilled from a larger Transformer architecture.^24,25^ The inputs to that model were the concatenated structured text strings of previous diagnoses and medical procedures, with the output being 256-dimensional embeddings. Using Principal Component Analysis (PCA), we reduced this to 60 components, retaining 88·9% of the variance, with a composite embedding score (mean of all components) used in the propensity score model.

We defined 24-hour mortality risk in the absence of major interventions as the outcome to be predicted by the study’s models. Major clinical interventions were defined as ICU admission, respiratory and ventilatory support, surgical operations, and anesthesia administration, within 24 hours following the initial NEWS recording. We defined the study’s target population as the entire population of adult patients with a hospital encounter exceeding 15 minutes. Since some patients receiving major rescuing interventions due to predicted high baseline risk, we applied inverse probability weighting (IPW) in our EWS validation to correct for intervention bias that would have distorted our results had we kept these patients in the evaluation (i.e. artifactual overestimation of mortality) or discarded them without weighting the remaining analytic cohort (i.e. selection bias). IPW allowed us to weight the patients without major interventions such that they approximated the overall target population, estimating the 24-hour mortality risk that would be observed without major interventions.^26^ This approach is referred to as “hypothetical strategy” in the novel predictimand framework. ^12^ To compute IPW, we estimated propensity scores for receiving major interventions using covariate balancing propensity scores, creating weights that made the analytic cohort of patients without major interventions representative of the overall target population (Appendix pp 4—6). We assessed covariate balance using standardized mean differences to evaluate whether patients without major interventions were representative of the overall hospitalized cohort.

We developed three logistic regression models in the final analytic cohort with different EWSs as predictors: NEWS, Simplified NEWS (NEWS without temperature and blood pressure parameters, removed as a potential pathway to reduce bedside measurement burden), and DEWS, i.e., Demographic Early Warning Score^4^: age and sex in addition to simplified vital sign scoring (Appendix pp 3, 6). We further developed a machine learning tree-based model, XGB-EWS, with all available predictors, including NEWS’s vital sign set plus diastolic blood pressure, laboratory values, number of previous encounters, history of ICU/respiratory support, clinical history embedding components, demographic variables (age, sex), and temporal features. All models incorporated the weights in both the model fitting and performance evaluation. A description of the models used can be found in the Appendix (pp 9—10).

Model performance was assessed on the basis of discrimination, calibration, and clinical utility using grouped ten-fold cross-validation with hospitals as clusters to account for hospital-level effects (Appendix p 2). Discrimination was measured using the AUC, i.e., the probability that the model assigned higher risks to patients who died than to those who survived. Calibration was assessed via the Brier score (mean squared difference between predicted probabilities and observed outcomes) and visual inspection of calibration plots. Clinical utility was evaluated using decision curve analysis, which compares the net benefit of using each model to guide clinical decisions against default strategies of treating all patients or treating no patients across different risk thresholds (Appendix pp 11, 29). We selected the risk thresholds as multiples of the weighted mortality prevalence (1×, 2×, 4×, 8×, 10×, 20×, and 30×), with subgroup analyses using subgroup-specific prevalences. An overview of the study’s methodology is presented in Figure 1.

**Figure 1:**
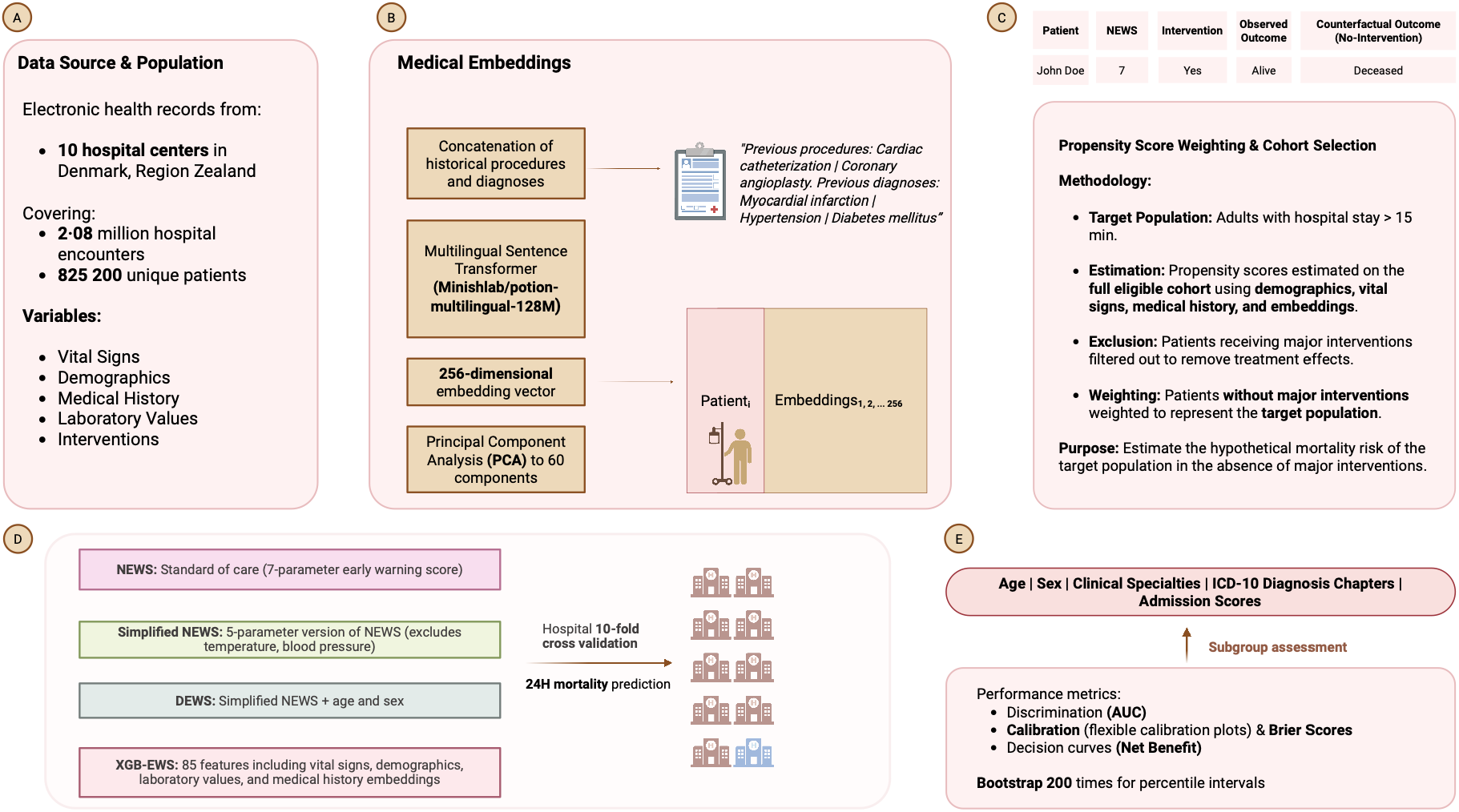
Overview of the study’s workflow. (A) Electronic health records from Danish hospitals capturing patient data, including vital signs, demographics, medical history, laboratory values, and clinical interventions. (B) To incorporate historical medical information, previous procedures and diagnoses were concatenated into text strings, then transformed into numerical embeddings using multilingual sentence transformers and dimensionality reduction. (C) Propensity score weighting created a balanced cohort that allows evaluation of early warning score performance in the counterfactual scenario where major interventions had not occurred. (D) Four prediction models were compared using hospital-clustered cross-validation to predict 24-hour mortality, ranging from the standard NEWS to machine learning approaches incorporating all available data. (E) Model performance was assessed through multiple metrics and subgroup analyses across patient demographics and clinical contexts.

The current work was approved by the hospital management and legal departments of the participating hospitals as a quality improvement project. According to Danish law, patient informed consent is not required for quality improvement projects. Therefore, approval from the regional scientific ethics committee was not required.

## Results

During the study period from October 16, 2018, to October 16, 2023, we identified 18 877 954 EWS measurements from 2 355 905 hospital encounters across 899 100 unique patients in 10 hospital networks. After excluding encounters with missing demographics and hospital stays of ≤15 minutes and restricting the analysis to the first NEWS assessment per hospitalization, the final cohort comprised 2 351 828 encounters of 898 662 individuals. We computed inverse probability weights using propensity scores estimated on the full cohort to account for major 24-hour interventions. Subsequently, we restricted the analysis to the 2 080 765 hospital encounters of 825 200 individuals without major interventions, applying the weights to make this analytical sample representative of the overall hospitalization cohort (Appendix, pp 23—25). Baseline characteristics of the weighted cohort are presented in Table 1. The weighted 24-hour mortality was 0·22% (4 741 deaths); median NEWS score (Interquartile Range) was 1 (2·00) overall, but (7·00 (5·00)) among patients who died within 24 hours. The weighted population comprised a majority (50·9%) of patients aged 18–65 years, with 50·4% female patients and 66·2% admitted to internal and acute medicine clinical specialties.

In the cross-validated analysis, XGB-EWS demonstrated the highest overall discrimination with an AUC of 0·935 (95% CI 0·930-0·938), followed by DEWS at 0·908 (95% CI 0·904-0·912), NEWS at 0·900 (95% CI 0·895-0·905), and Simplified NEWS at 0·878 (95% CI 0·871-0·883). The hospital-specific discriminative performance of the models (Figure 2) showed consistent patterns across the strata. Model calibration was assessed using flexible calibration curves with restricted cubic splines. For the overall population, all models demonstrated reasonable calibration across the risk spectrum (Figure 3). When stratified by age group, all models except DEWS showed a pattern of risk overestimation in the youngest group (18-65 years), adequate calibration in the middle age group (66-80 years), and underestimation in the oldest group (≥80 years). Sex-stratified analysis revealed similar calibration performance across all models. Hospital-specific Brier scores and detailed subgroup calibration analyses are provided in the Appendix (pp 26—28). The feature importance analysis of XGB-EWS using SHAP values indicated that vital signs dominated predictions despite inclusion of other metadata (Appendix, p 30).

**Figure 2:**
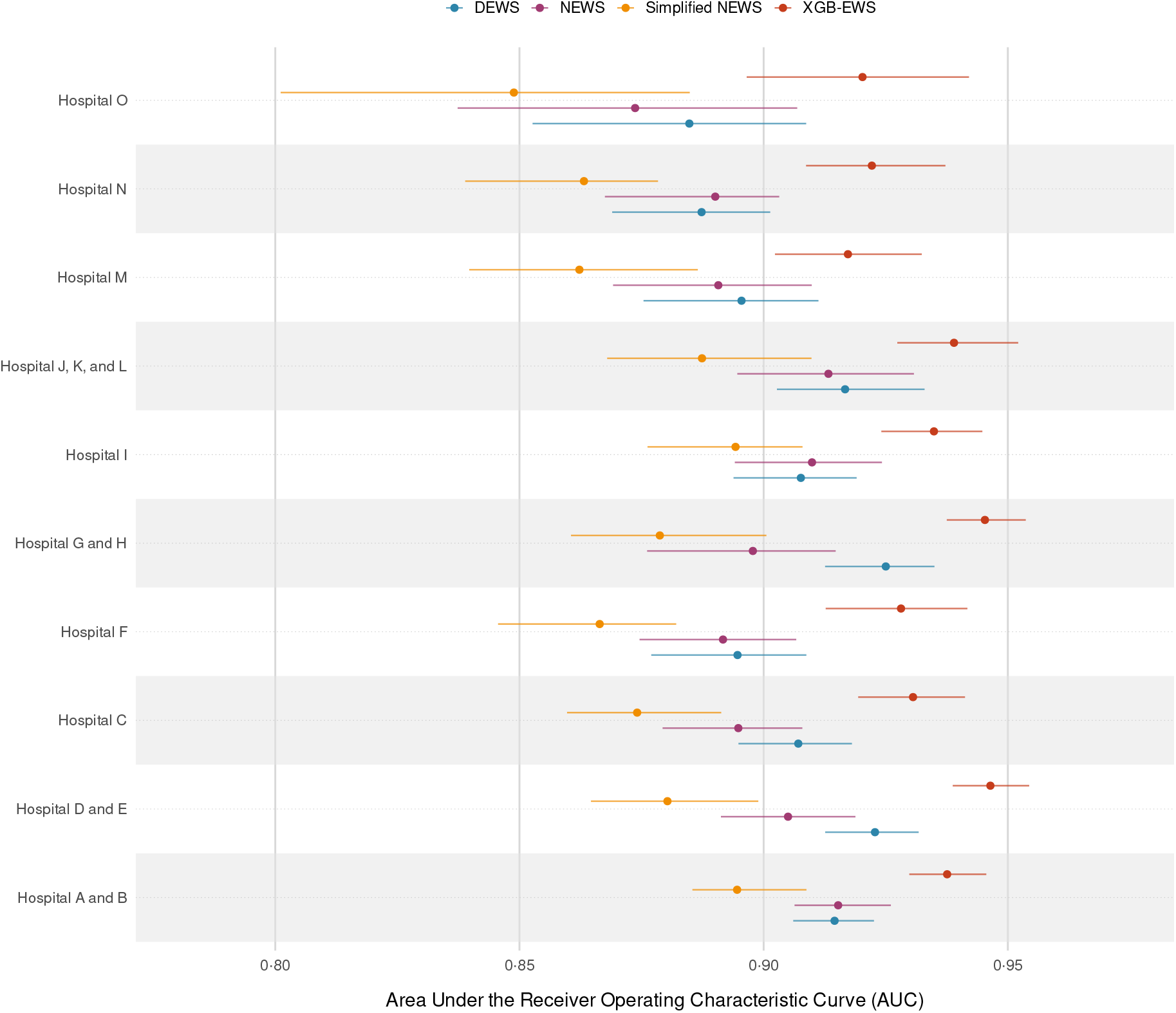
Forest plot of hospital-specific discriminative performance of EWS algorithms. The Area under the Receiver Operating Characteristic Curve (AUC) with 95% confidence intervals for predicting 24-hour mortality across the 10 hospital clusters is illustrated. Each point represents the discriminative performance of the four prediction models within individual hospital settings. Confidence intervals were calculated using 200 bootstrap resamples with the percentile method (2·5th and 97·5th percentiles). The four models were XGB-EWS, DEWS, NEWS, and Simplified NEWS, and evaluated across all 15 participating hospitals in the study’s hospital network.

**Figure 3:**
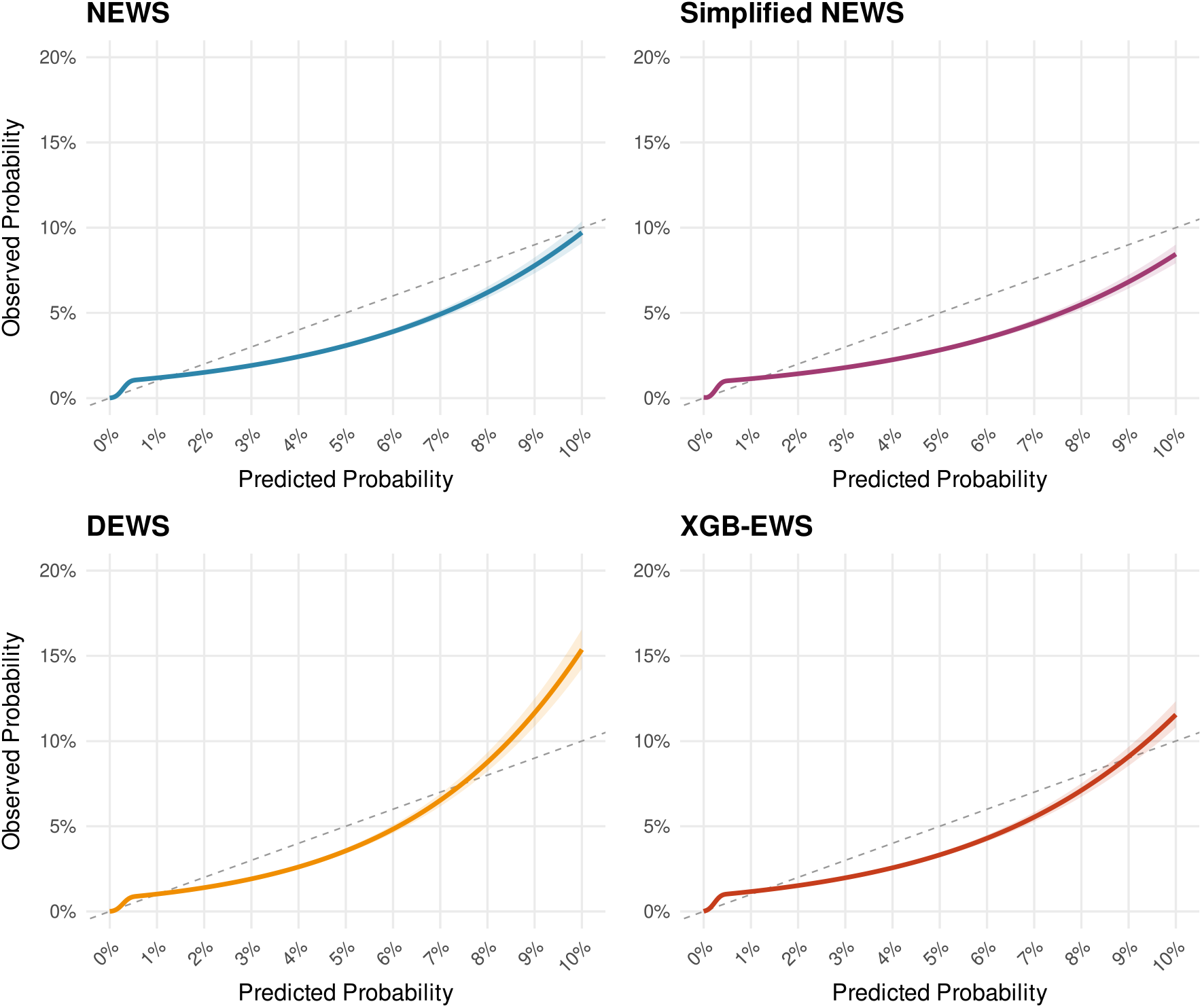
Model calibration across the overall population. Flexible calibration plots comparing predicted probabilities versus observed 24-hour mortality risks for the four evaluated models (NEWS, Simplified NEWS, DEWS, and XGB-EWS). The curves were estimated using logistic regression with restricted cubic splines (4 knots) to visualize calibration performance continuously across the risk spectrum. The solid-colored lines represent the estimated calibration curves, while the surrounding shaded areas indicate the 95% confidence intervals. The dashed gray diagonal line represents perfect calibration (where predicted risk equals observed risk). All estimates were calculated using inverse probability weights to correct for intervention bias and reflect the target population.

We conducted decision curve analysis to evaluate clinical utility at thresholds corresponding to NEWS escalation protocols (Appendix, p 11). Table 2 shows the Net Benefit and corresponding differences of the study’s models. At the medium-risk threshold of 0·0088 (corresponding to NEWS 5-6, where urgent ward-based response is typically triggered), XGB-EWS showed 1·3 additional correct identifications per 10000 patients compared to NEWS, while Simplified NEWS showed 1·5 fewer identifications than NEWS. At the high-risk threshold of 0·0441 (corresponding to NEWS ≥7, where emergency response is mandated), these differences were 1·8 additional for XGB-EWS versus NEWS and 1·1 fewer for Simplified NEWS versus NEWS. The maximum net benefit differences across all evaluated thresholds were 1·9 per 10000 patients between XGB-EWS and NEWS, and 1·5 per 10000 between NEWS and Simplified NEWS. We provide a full decision curve for all models and the overall population in the Appendix (p 29). However, when stratified by NEWS thresholds (Figure 4), net benefit differences varied substantially by baseline risk. In the high-risk stratum (NEWS ≥7), XGB-EWS achieved a maximum gain of 78·1 per 10000 compared to NEWS, while Simplified NEWS showed a maximum loss of 60·2 per 10000 relative to NEWS. In the medium-risk stratum (NEWS 5-6), maximum differences were 31·7 per 10,000 for XGB-EWS versus NEWS and 16·7 per 10000 for NEWS versus Simplified NEWS. These differences progressively diminished in lower-risk strata. In the most prevalent diagnostic groups (symptoms and signs, R-codes, 26·4% of encounters; factors influencing health 11·0%; circulatory 10·4%; injury or poisoning 9·5%; respiratory 6·6%), net benefit differences were modest (XGB-EWS vs NEWS: 1·7 to 5·3 per 10000; NEWS vs Simplified NEWS: 0·8 to 3·1 per 10000). DEWS showed gains over NEWS in respiratory and mental diagnoses, with mixed results in circulatory ones but not in R-codes, the largest group, where the two performed comparably. Larger net benefit differences were observed in patients aged 80 years and above (18·5% of the cohort) and in low-prevalence diagnostic subgroups such as blood-related diseases (1·0%) (Appendix, pp 12–18). Sensitivity analyses using 48-hour and 72-hour mortality endpoints, a composite outcome of ICU admission or 24-hour mortality, and restricting to hospital stays >60 minutes showed consistent patterns, with maximum net benefit differences ranging from 1·5 to 7·0 per 10000 patients across endpoints (Appendix, pp 19—22).

**Figure 4:**
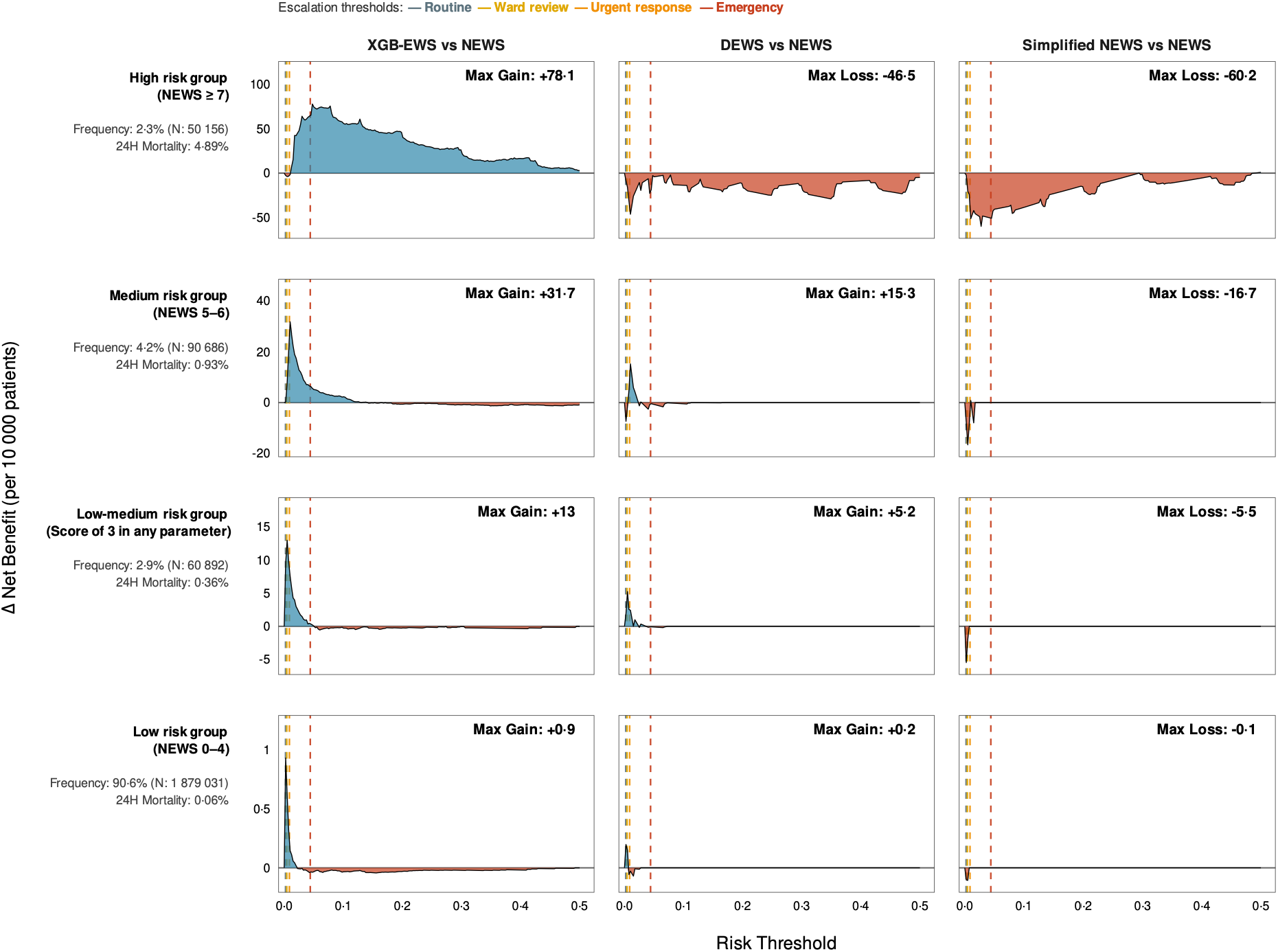
Net benefit differences between prediction models across clinical risk groups. The panel grid displays the difference in Net Benefit (per 10 000 patients) between each evaluated model and the standard NEWS reference. Rows represent patient subgroups stratified by their initial NEWS score: High (NEWS >= 7), Medium (NEWS 5–6), Low-Medium (single parameter score of 3), and Low (NEWS 0–4). Columns represent the comparison of the standard NEWS against XGB-EWS (machine learning model), DEWS (Simplified NEWS with age/sex), and Simplified NEWS (NEWS without blood pressure/temperature). Blue-shaded areas indicate probability thresholds at which the alternative model provides superior clinical utility compared to NEWS (Gain). Red-shaded areas indicate thresholds at which the alternative model has inferior utility (Loss). The curves are plotted across a continuous probability range (0·00 to 0·50). Vertical dashed lines indicate the four NEWS-based clinical escalation thresholds (Routine, Ward review, Urgent response, Emergency) at their corresponding risk thresholds. Note that net benefit differences represent additional correct mortality identifications, not direct mortality reductions; clinical outcomes depend on how EWS scores are used in clinical decisions, subsequent care quality, treatment availability, patient factors beyond the model’s reach, and the severity of illness.

## Discussion

This validation study of 2·08 million hospital encounters enabled detailed evaluation of EWS performance across diverse clinical populations and subgroups. We found that our gradient boosting tree-based model achieved better discriminative performance compared to traditional scoring systems, consistent with previous research.^7,9,27,28^ However, decision curve analysis, an assessment rarely performed in EWS studies^1^, yielded a more nuanced picture. Our predominantly low-acuity population, with low overall mortality, allowed all models to effectively discriminate between extremes, yielding high AUC values consistent with prior reports in broad hospital populations.^16^ Nevertheless, stratification by NEWS thresholds revealed that this uniformly high discrimination masked the heterogeneity in clinical utility across categories. In high-risk patients requiring emergency response protocols, both the machine learning approach and the full NEWS score provided considerably larger incremental benefits compared to simplified approaches, while in low-risk populations under standard monitoring, these differences became minimal. This demonstrates that better discrimination metrics do not uniformly translate into clinical benefit, with model complexity potentially providing value in high-risk escalation categories.

Sensitivity analyses extending the prediction horizon to 48 and 72 hours showed a modest increase in the machine learning advantage over time, suggesting that vital signs dominate immediate deterioration prediction, whereas laboratory values and history may contribute more to medium-term prognostication. In the overall population, Simplified NEWS remained close to NEWS across all time horizons and for the composite outcome of ICU admission or mortality, suggesting limited average loss from removing blood pressure and temperature. However, this population-level pattern masked clinically important heterogeneity. Decision curve analysis showed that the advantage of XGB-EWS and full NEWS over Simplified NEWS became more pronounced in high-risk subpopulations, indicating that simplification may be less appropriate when baseline risk is elevated. A related form of heterogeneity emerged across diagnostic groups: the added value of demographic information over vital signs was concentrated in patients with established organ-system diagnoses, where age and sex modify disease-specific mortality. It was absent in undifferentiated R-coded presentations, where heterogeneous pathophysiologies leave vital signs to carry the predictive signal. Taken together, these subgroup findings support baseline stratification with NEWS and generate a hypothesis for prospective cluster-randomised trials: lower-risk patients could be monitored with Simplified NEWS, supplemented by clinical judgement, whereas higher-risk patients or those of clinical concern could continue with NEWS or more enriched assessment. This workflow extends the I-EWS approach^6^, in which clinicians may adjust the calculated score based on clinical assessment, by applying this principle to a simplified vital-signs base. From a practical standpoint, implementation of Simplified NEWS would require minimal retraining given its structural similarity to NEWS, while XGB-EWS serves as proof-of-concept for settings with integrated electronic health records.

Our study has several limitations. First, implementing advanced models like XGB-EWS requires robust IT infrastructure for real-time data ingestion, unlike standard scoring systems that function with manual calculation, creating potential deployment barriers in resource-limited settings. Second, the retrospective observational design of the study limits causal inference about the effectiveness of the proposed models, and the Danish healthcare context may limit generalizability to other healthcare systems with different organizational structures or data availability. In particular, ethnicity is not routinely recorded in Danish electronic health records and was therefore unavailable for our analyses, which may limit generalisability to more ethnically diverse populations. Third, despite our propensity score weighting approach incorporating vital signs, demographics, medical history embeddings, and hospital-specific factors, unmeasured variables, such as clinical intuition, real-time resource constraints, or rapid physiological changes, may persist, potentially biasing our hypothetical risk estimates. Additionally, our definition of major clinical interventions, while clinically informed, could be refined in future studies to explore sensitivity to different criteria. For out-of-hospital deaths, the recorded time of death reflects the certifying physician’s estimate at post-mortem examination, which may introduce minor misclassification at the 24-hour boundary. This effect is likely limited because only boundary cases are affected and all models use the same outcome definition. Most importantly, while our study demonstrates comparable predictive performance and quantifies potential efficiency gains, clustered randomized controlled trials are needed to evaluate real-world implementation effectiveness. However, our study has several strengths that advance the field. Methodologically, we applied the predictimand framework using propensity score weighting to mitigate intervention bias, allowing a model-based approximation of performance in the absence of interventions. The scale of 2·08 million encounters across 10 hospital clusters enabled detection of clinically meaningful differences across diverse subgroups and institutional settings. Finally, by quantifying net benefit across approaches, we provide a basis for weighing predictive gains of machine learning against the NEWS system and its variants.

In conclusion, our findings suggest that healthcare systems should weigh predictive performance against workflow optimization. While simplified systems achieve comparable performance at the population level, more advanced models may offer greater clinical utility in high-risk subpopulations. These results support prospective evaluation of stratified implementation strategies based on baseline risk at the first NEWS assessment.

## Data Availability

Access to the dataset supporting the conclusion of this article can only legally be accessed on acceptance from the Danish Data Protection Agency, the government body that regulates the use of Danish registry data. Interested parties can contact (PBN & KKI) for further information. We confirm that others, given a suitable project judged by the Danish Data Protection Agency, would be able to access these data in the same manner as the authors. We also confirm that the authors do not have any special access privileges that others do not have. The codebase for the analysis of the current study can be found online: https://github.com/MLGlobalHealth/Early_Warning_Scores

## Author’s contributions

AK, NS, KKI, PBN, and TL conceptualized and designed the study. SB provided supervision for the project. AK and NS conducted all quantitative analyses. AK and SB wrote the first draft of the manuscript. AK, NS, KKI, and PBN verified the raw data. All authors provided methodological insights on the manuscript, had full access to all the data, seen and approved the final text, and were involved in revising the manuscript. All authors had final responsibility for the decision to submit for publication.

## Acknowledgments

This work is funded from the Novo Nordisk Foundation via The Novo Nordisk Young Investigator Award (NNF20OC0059309). AK acknowledges funding from the Novo Nordisk Foundation via The Novo Nordisk Young Investigator Award (NNF20OC0059309). NS and MPK acknowledge funding from The Danish National Research Foundation via a chair grant (DNRF160). TVV is supported by the “Data Science Investigator - Emerging 2022” grant from the Novo Nordisk Foundation (NNF22OC0075284). AGZ and The Copenhagen Health Complexity Center are funded by TrygFonden. SB acknowledges funding from the MRC Centre for Global Infectious Disease Analysis (reference MR/X020258/1). SB acknowledges support from the Novo Nordisk Foundation via The Novo Nordisk Young Investigator Award (NNF20OC0059309). SB acknowledges the Danish National Research Foundation (DNRF160) through the chair grant. SB acknowledges support from Schmidt Sciences via the Schmidt Polymath Award (G-22-63345). We acknowledge the use of BioRender for the creation of Figure 1 (Katsiferis, A. (2025) https://BioRender.com/f1b7o4s.

