## Supplementary material for "Development and validation of early warning scores predicting 24-hour mortality addressing intercurrent medical interventions: a multi-centre retrospective cohort study of 2·08 million hospital encounters": Tables 1 & 2

**Table 1: Baseline characteristics of untreated weighted patients by 24-Hour mortality status**

| Characteristic | Overall<br>N = 2 080 765 | Deceased<br>N = 4 741 | Alive<br>N = 2 076 024 |
|---|---|---|---|
| **Age group** | | | |
| 18-65 | 1 053 140 (50·9%) | 542 (11·6%) | 1 052 598 (51·0%) |
| 66-80 | 629 221 (30·6%) | 1 721 (36·9%) | 627 500 (30·6%) |
| 80+ | 398 404 (18·5%) | 2 478 (51·4%) | 395 926 (18·5%) |
| **Sex** | | | |
| Female | 1 043 017 (50·4%) | 2 114 (44·8%) | 1 040 903 (50·4%) |
| Male | 1 037 748 (49·6%) | 2 627 (55·2%) | 1 035 121 (49·6%) |
| **Previous hospitalizations (count)** | 1·0 (0·0 - 3·0) | 2·0 (1·0 - 5·0) | 1·0 (0·0 - 3·0) |
| **Hospital** | | | |
| Hospital D and E | 251 472 (11·6%) | 522 (10·7%) | 250 950 (11·6%) |
| Hospital G and H | 230 579 (11·0%) | 382 (8·0%) | 230 197 (11·0%) |
| Hospital O | 41 096 (2·0%) | 129 (2·7%) | 40 967 (2·0%) |
| Hospital A and B | 440 266 (21·2%) | 1 028 (21·7%) | 439 238 (21·2%) |
| Hospital M | 131 665 (6·1%) | 331 (6·8%) | 131 334 (6·1%) |
| Hospital J, K, and L | 159 428 (7·6%) | 389 (8·2%) | 159 039 (7·6%) |
| Hospital C | 249 334 (12·0%) | 603 (12·7%) | 248 731 (12·0%) |
| Hospital N | 114 399 (5·4%) | 400 (8·3%) | 113 999 (5·4%) |
| Hospital F | 238 829 (12·0%) | 391 (8·5%) | 238 438 (12·0%) |
| Hospital I | 223 697 (11·1%) | 566 (12·3%) | 223 131 (11·1%) |
| **Clinical specialty** | | | |
| Cardiopulmonary | 151 050 (7·3%) | 230 (4·9%) | 150 820 (7·3%) |
| Critical Care | 64 262 (3·1%) | 40 (0·9%) | 64 222 (3·2%) |
| Gastroenterology | 20 518 (1·0%) | 27 (0·6%) | 20 491 (1·0%) |
| Internal & Acute Medicine | 1 382 512 (66·2%) | 3 840 (80·7%) | 1 378 672 (66·1%) |
| Nephrology | 15 055 (0·7%) | 21 (0·4%) | 15 034 (0·7%) |
| Neurology | 54 715 (2·7%) | 97 (2·2%) | 54 618 (2·7%) |
| Oncology & Hematology | 48 048 (2·3%) | 101 (2·1%) | 47 947 (2·3%) |
| Surgical | 303 709 (14·7%) | 347 (7·3%) | 303 362 (14·7%) |
| Other | 40 896 (2·0%) | 38 (0·8%) | 40 858 (2·0%) |
| **Consciousness** | | | |
| A | 2 064 146 (99·2%) | 3 626 (75·7%) | 2 060 520 (99·2%) |
| VPU | 16 619 (0·8%) | 1 115 (24·3%) | 15 504 (0·8%) |
| **Supplemental oxygen** | | | |
| Air | 1 860 028 (89·6%) | 1 729 (36·9%) | 1 858 299 (89·7%) |
| Oxygen | 220 737 (10·4%) | 3 012 (63·1%) | 217 725 (10·3%) |
| **Respiratory rate (breaths/min)** | 16·0 (16·0 - 18·0) | 21·0 (18·0 - 28·0) | 16·0 (16·0 - 18·0) |
| **Pulse (beats/min)** | 79·0 (68·0 - 90·0) | 93·0 (77·0 - 110·0) | 79·0 (68·0 - 90·0) |
| **Temperature (°C)** | 36·7 (36·4 - 37·1) | 36·6 (36·1 - 37·2) | 36·7 (36·4 - 37·1) |
| **Oxygen saturation (%)** | 97·0 (96·0 - 99·0) | 95·0 (91·0 - 97·0) | 97·0 (96·0 - 99·0) |
| **Systolic blood pressure (mm Hg)** | 133·0 (119·0 - 149·0) | 116·0 (96·0 - 138·0) | 133·0 (119·0 - 149·0) |
| **Diastolic blood pressure (mm Hg)** | 76·0 (67·0 - 86·0) | 68·0 (56·0 - 81·0) | 76·0 (67·0 - 86·0) |
| **Previous ICU/respiratory support** | | | |
| No history of ICU or respiratory support | 2 046 815 (98·4%) | 4 606 (97·2%) | 2 042 209 (98·4%) |
| Previous history of ICU or respiratory support | 33 950 (1·6%) | 135 (2·8%) | 33 815 (1·6%) |
| **NEWS score** | 1·0 (0·0 - 2·0) | 7·0 (4·0 - 9·0) | 1·0 (0·0 - 2·0) |
| **Hemoglobin (mmol/L)** | 8·3 (7·5 - 9·0) | 7·6 (6·7 - 8·4) | 8·3 (7·5 - 9·0) |
| **Leukocytes (×10⁹/L)** | 7·6 (6·3 - 9·4) | 8·3 (6·8 - 10·3) | 7·6 (6·3 - 9·4) |
| **Platelets (×10⁹/L)** | 250·0 (206·0 - 300·5) | 245·0 (194·0 - 308·0) | 250·0 (206·0 - 300·5) |
| **Creatinine (µmol/L)** | 74·0 (63·0 - 90·0) | 82·5 (65·0 - 110·0) | 74·0 (63·0 - 90·0) |
| **Alanine aminotransferase (U/L)** | 23·0 (17·5 - 32·0) | 20·0 (15·0 - 28·0) | 23·0 (17·5 - 32·0) |
| **Lactate dehydrogenase (U/L)** | 192·0 (170·0 - 222·0) | 210·0 (180·0 - 250·0) | 192·0 (170·0 - 222·0) |
| **Albumin (g/L)** | 36·0 (32·0 - 39·0) | 32·0 (28·0 - 35·0) | 36·0 (32·0 - 39·0) |
| **C-reactive protein (mg/L)** | 5·0 (1·4 - 21·0) | 17·0 (5·2 - 45·0) | 5·0 (1·4 - 21·0) |
| **Lactate, arterial blood (mmol/L)** | 1·2 (0·9 - 1·8) | 1·5 (1·0 - 2·4) | 1·2 (0·9 - 1·8) |
| **Troponin T (ng/L)** | 19·5 (8·0 - 57·0) | 48·0 (23·0 - 106·0) | 19·5 (8·0 - 57·0) |
| **Lactate, venous blood (mmol/L)** | 1·4 (1·0 - 2·0) | 1·5 (1·1 - 2·2) | 1·4 (1·0 - 2·0) |

Values are Raw N (Weighted %) or Weighted Median (IQR). N represents the unweighted sample size. Percentages and medians are weighted to represent the target population. IQR = (25th - 75th percentile).

**Table 2: Net benefit comparison of different strategies and models at specified risk thresholds**

| | Net Benefit per 10 000 Patients | | | | | | Net Benefit Difference per 10 000 Patients | | | |
|---|---|---|---|---|---|---|---|---|---|---|
| **Risk Threshold** | **Treat All** | **Treat None** | **NEWS** | **Simplified NEWS** | **DEWS** | **XGB-EWS** | **NEWS - XGB-EWS** | **NEWS - Simplified NEWS** | **DEWS - XGB-EWS** | **NEWS - DEWS** |
| 0·0022 | 0·0 | 0·0 | 14·6 | 14·1 | 14·4 | 15·7 | -1·1 | 0·5 | -1·3 | 0·2 |
| 0·0044 | -22·1 | 0·0 | 12·3 | 11·5 | 12·2 | 13·5 | -1·2 | 0·8 | -1·3 | 0·1 |
| 0·0088 | -66·7 | 0·0 | 9·9 | 8·4 | 9·4 | 11·2 | -1·3 | 1·5 | -1·8 | 0·5 |
| 0·0176 | -157·1 | 0·0 | 6·9 | 5·9 | 6·6 | 8·8 | -1·9 | 1·0 | -2·2 | 0·3 |
| 0·0220 | -202·9 | 0·0 | 6·3 | 5·2 | 6·0 | 8·1 | -1·8 | 1·1 | -2·1 | 0·3 |
| 0·0441 | -438·1 | 0·0 | 4·1 | 3·0 | 3·6 | 5·9 | -1·8 | 1·1 | -2·3 | 0·5 |
| 0·0661 | -684·5 | 0·0 | 2·9 | 2·0 | 2·7 | 4·7 | -1·8 | 0·9 | -2·0 | 0·2 |

Positive values of net benefit difference indicate the first model has higher net benefit; negative values indicate the comparator is better. Risk thresholds are chosen as multiples of the mortality prevalence in the final analytic cohort (N = 2 080 765)
