## Supplementary Appendix for "Development and validation of early warning scores predicting 24-hour mortality addressing intercurrent medical interventions: a multi-centre retrospective cohort study of 2·08 million hospital encounters"

### Supplemental Content 1 – Additional Details

#### Verification of mortality timestamps

In Denmark, the electronic death certificate, completed by the attending physician at the time of post-mortem examination, records both the date and the time of death with minute-level precision. The certificate is submitted electronically through the Danish Health Data Authority's reporting system and propagated to downstream systems, including Sundhedsplatformen, the regional electronic health record system serving the Capital Region of Denmark and Region Zealand, from which all of our analytical data were drawn. Sundhedsplatformen therefore carries the time-of-death timestamps with minute-level resolution.

For deaths occurring during an index hospital encounter, the time of death is documented directly in Sundhedsplatformen by attending clinical staff at the bedside, providing real-time minute-level precision. For deaths occurring after discharge of the index encounter, the recorded time reflects the certifying physician's assessment at post-mortem examination, which may be an estimate (for example, when the death was unwitnessed) rather than a directly observed event, and therefore constitutes a potential source of minor misclassification at the 24-hour boundary.

To verify whether the timestamps used to define 24-hour outcome reflect authentic clinical entries rather than defaults from missing time-of-day information, we examined below their time-of-day distribution (Figure 1): authentic timestamps should be distributed across the day, whereas systematic defaulting would manifest as concentration at one specific time. Of the 4 741 deaths within 24 hours of the initial NEWS recording, 4 579 (96·6%) occurred during the index hospital encounter with clinician-recorded timestamps, and 162 (3·4%) occurred after discharge. Because any potential misclassification at the 24-hour boundary would affect only boundary cases and arise from the shared outcome definition used by all four models (NEWS, Simplified NEWS, DEWS, and XGB-EWS), its impact on between-model comparisons is expected to be limited.

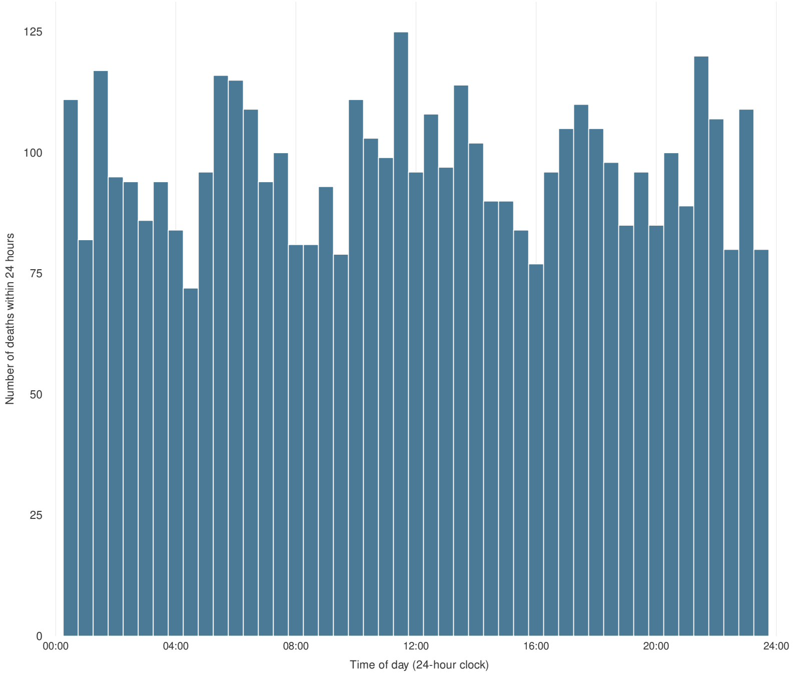

**Figure 1.**
Time-of-day distribution of deaths within 24 hours of the initial NEWS recording. Histogram of all deaths within the 24-hour outcome window in the analytical cohort (n = 4 741), binned in 30-minute intervals.

#### NEWS Implementation in the study's hospitals

Per regional clinical guidelines, NEWS monitoring is mandated for all adult patients (≥18 years) upon receiving admitted status across somatic hospitals and clinical specialties. Patients presenting to emergency departments are initially assessed using triage protocols; NEWS scoring begins when admission occurs. The seven vital sign parameters (respiratory rate, oxygen saturation, supplemental oxygen, systolic blood pressure, pulse, consciousness, and temperature) are measured at intervals determined by the patient's current NEWS score.

#### Handling of Missing Data

Median (continuous variables) and mode (categorical) imputation were applied only where complete input matrices were required for model fitting or preprocessing. Missing values in clinical history text fields (SKS descriptions) were preserved as missingness indicators and handled implicitly through the text processing pipeline. Vital signs and demographic variables demonstrated missingness of <0·5%. The XGB-EWS model used XGBoost's native 'sparsity-aware split finding' capability to handle missing laboratory data.[1] For the latter, which exhibited high missingness rates ranging from 26·3% (Creatinine) to 93·0% (Troponin T), explicit imputation was avoided. Instead, during the training phase, the model learned an optimal 'default direction' (left or right child node) for missing values at each tree split to minimize the training loss. This methodological approach allows the model to capture informative missingness, i.e., a phenomenon where the clinical decision not to order a specific test (e.g., the absence of an arterial lactate measurement) contains predictive information regarding patient acuity that would be obscured by median imputation.

#### Grouped Cross-Validation Strategy

The 15 participating physical hospital locations were organized into 10 clusters corresponding to the actual merged hospital entities (as detailed in **Table 1** of the main manuscript). For example, Hospital D and E were merged administratively on January 1, 2012. These 10 hospital clusters cover the Capital Region and the Zealand Region, which together represent 2·76 million people, accounting for almost 46% of the entire Danish population. The two regions encompass both densely populated areas (Copenhagen metropolitan area) and less densely populated communities in Zealand. The 15 hospitals were selected based on existing data-sharing collaborations (common employment of the *Sundhedsplatformen* record system) that enabled access to comprehensive electronic health records with the granular data required for our analysis. The study period was selected to capture a data volume sufficient for meaningful analyses while incorporating potential temporal variation.

To evaluate the generalizability of the models and simulate their performance in unseen clinical environments, we employed a **Grouped 10-Fold Cross-Validation** strategy (also known as "Leave-One-Hospital-Group-Out" validation). Unlike standard k-fold cross-validation, which randomly splits the data and can lead to data leakage in which the model learns site-specific idiosyncrasies, our approach ensured that the training and validation sets were strictly separated by hospital location.

The validation process proceeded as follows:

**Clustering:** Data was partitioned into 10 distinct folds corresponding to the 10 merged hospital entities/clusters.
**Iteration:** In each of the 10 iterations, one hospital entity was held out as the independent test set, while the remaining 9 entities were combined to form the training set.
**Training:** The models were trained on the pooled data from the 9 training entities.
**Evaluation:** The trained models were applied to the held-out hospital entity to generate predicted probabilities.
**Aggregation:** This cycle was repeated until every encounter in the dataset was assigned a predicted probability derived from a model that had never seen data from that specific hospital entity during training.
This method assesses the model's ability to generalize across different institutions with varying local populations and operational practices.

#### Sample Size Considerations

We performed sample size calculations using the *pmsampsize* package to assess adequacy for prediction model development.[2] Based on an anticipated AUC of 0·8, 100 predictor parameters for the most complex model, and an observed 24-hour mortality prevalence of 0·22%, the minimum required sample size was 289 823 encounters with 638 mortality events to achieve an events-per-parameter ratio of 6·38.

#### Ablation Analysis & Simplified NEWS

To address the priority of workflow optimisation in routine hospital monitoring, we sought a simplified version of NEWS that removes its most time-consuming components (blood pressure and temperature), which represent the most plausible workflow lever available in current clinical practice. To assess whether this simplification carried an acceptable predictive cost**,** we performed a complete ablation analysis based on the full NEWS score (currently used in the Danish hospitals included in the study). Specifically, we evaluated the increase in the prediction error (as measured by the Brier Score) when each component was removed from the score. The results are presented below in Figure 2. We observed that the 95% confidence intervals for the prediction error of 24h mortality overlapped with those of the full NEWS for all components evaluated. This overlap was the second smallest for systolic blood pressure (with only consciousness producing a smaller overlap) but remained present, satisfying our pre-specified data-centric criterion for removal. The resulting model, Simplified NEWS, was therefore taken forward as a candidate, allowing us to test the hypothesis that a less operationally intensive scoring model can offer comparable predictive capacity to more complex scoring systems for detecting clinical deterioration in patients.

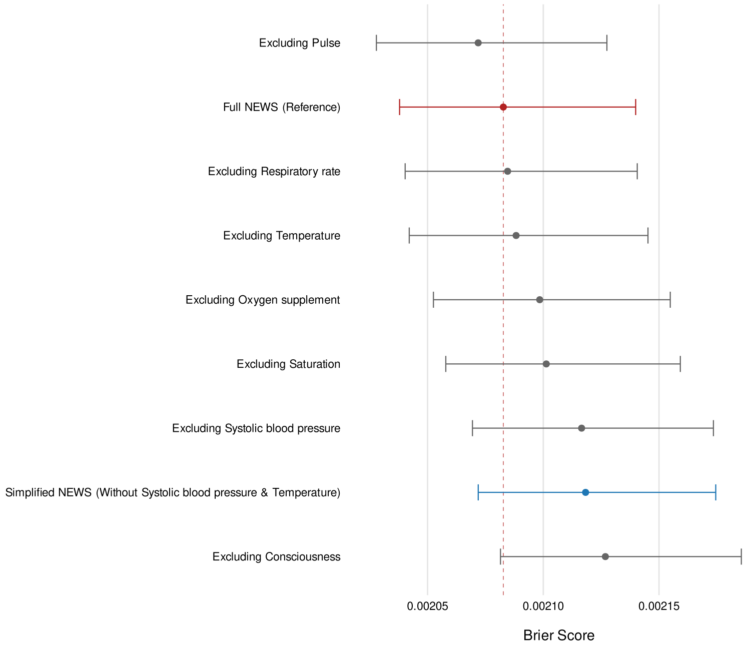

**Figure 2.**

Ablation analysis of the marginal contribution of National Early Warning Score (NEWS) parameters to predictive performance. The forest plot illustrates the Brier Score (Mean Squared Error) for 24-hour mortality prediction in the final analytic cohort (N = 2 080 765). The vertical dashed red line represents the reference performance of the Full NEWS model. Each circle represents the mean Brier Score calculated across 200 bootstrap resamples for a model with that specific parameter removed. Lower Brier Scores indicate better model accuracy and calibration. Horizontal bars denote 95% confidence intervals derived from the bootstrap distribution.

#### Temporal Distribution & Repeat Visit Patterns

Our pre-exclusion cohort (including individuals exposed to major interventions and those who did not) included 898 662 unique patients contributing 2 351 828 hospital encounters over five years (2018-2023). We analyzed temporal patterns in both the pre-exclusion dataset and our final analytical cohort (n = 2 080 765 after excluding those with major interventions).

**Repeat visit distribution:** In the final analytical cohort, 36·7% of encounters were first visits during the study period (previous hospitalizations = 0), while 63·3% were repeat visits (previous hospitalizations ≥1), with a median of 1 prior hospitalization (IQR: 0-3, mean: 3·0) (**Table 1**).

**Temporal spacing:** Among encounters with a prior visit in the pre-exclusion dataset (n = 1 453 166), 34·2% occurred within 24 hours of the previous encounter (**Table 2**). This clustering primarily reflects intrahospital transfers (31·0% with overlapping administrative records where a new encounter was recorded before the previous discharge was finalized) rather than independent re-encounters. For non-overlapping encounters (n = 1 003 315), median latency was 107 days (IQR: 19·7-363 days).

**Table 1: Previous Hospitalizations in Final Analytical Cohort (n = 2 080 765)**

| Previous Hospitalizations | N | % | Summary Statistics | Value |
|---|---|---|---|---|
| 0 (First visit) | 764 106 | 36·7 | Median | 1·0 |
| ≥1 (Repeat visit) | 1 316 659 | 63·3 | IQR | 0-3 |
| | | | Mean | 3·0 |
| Previous hospitalizations = count of prior encounters during study period (2018-2023) at time of index encounter. | | | | |

We retained all encounters rather than excluding short-latency visits because: (1) exclusion would remove 34·2% of repeat encounters and bias toward simpler-case patients; (2) intrahospital transfers represent clinical contexts where early warning scores are specifically designed to function; and (3) retaining repeat encounters better reflects operational use of EWSs in routine care. Hospital-clustered cross-validation was used to assess transportability across hospital entities, and previous hospitalizations were included in the propensity score model to account for prior utilization history. These steps were intended to preserve the operational encounter-level target of prediction rather than to model patient-level correlation directly.

**Table 2: Latency Between Consecutive Encounters (Pre-Exclusion Cohort)**

| Latency Category | N | % |
|---|---|---|
| Overlapping (negative) | 449 851 | 31·0 |
| Same-day (0-24h) | 45 844 | 3·2 |
| 1-7 days | 103 325 | 7·1 |
| 8-30 days | 157 212 | 10·8 |
| 31-90 days | 164 592 | 11·3 |
| 90+ days | 532 342 | 36·6 |
| **Total with prior visit** | **1 453 166** | **100·0** |

#### Methodological Handling of Intervention Bias

##### *Rationale and Predictimand Framework*

Standard retrospective validation of Early Warning Scores (EWS) is susceptible to intervention bias. In clinical practice, patients with high risk scores often receive immediate life-saving interventions (e.g., ICU admission, mechanical ventilation), which modifies their mortality outcome. If these patients are included in the training data with their observed outcomes (survival), the model may paradoxically learn to associate high-risk vital signs with lower mortality. Conversely, excluding these high-risk patients introduces selection bias, shifting the distribution of the training data toward lower-acuity presentations.

To address this, we adopted a hypothetical strategy within the predictimand framework.[3] We sought to estimate the risk of 24-hour mortality for the entire target population under the counterfactual condition that major interventions had *not* occurred.

##### *Inverse Probability Weighting (IPW) Methodology*

We utilized Inverse Probability Weighting (IPW) to construct a synthetic cohort. This process involved 5 steps:

1. **Propensity Score Estimation:** We estimated the propensity score, *e(X)*, defined as the probability of receiving a major intervention (A=1) given a set of baseline covariates (X): *e(X)* = P(*A* = 1 | *X*), using the full eligible cohort. The propensity model was fitted using the Covariate Balancing Propensity Score (CBPS) method to maximize balance. The covariate vector (X) included age, sex, number of previous hospitalisations, hospital site, full vital sign parameters, history of ICU/respiratory support, and a composite embedding score derived from natural language processing of clinical history to capture underlying comorbidity burden.
2. **Exclusion of Intervened Patients:** We consequently excluded patients who actually received major interventions (A= 1) from the outcome modeling, as their observed mortality outcomes do not reflect the natural trajectory of deterioration under standard ward care.
3. **Reweighting the Untreated:** We applied weights to the remaining untreated patients (A = 0) to make them representative of the total target population. The weight w for each retained patient was calculated as the inverse probability of not receiving the intervention: *w* = 1 / (1 − *e(X)*). Consequently, untreated patients with a high probability of intervention (i.e., high acuity) were up-weighted to represent the high-risk patients who were excluded.
4. **Model Training Integration:** These calculated weights were explicitly incorporated into the training phase of all predictive models. For the logistic regression models (NEWS, Simplified NEWS, DEWS), weights were used to maximize the weighted likelihood. For the machine learning model (XGB-EWS), weights were incorporated into the objective loss function during tree construction. This ensures that all models minimize prediction error relative to the target population's risk profile, rather than the observed population.
5. **Weighted Performance Evaluation:** All performance metrics were calculated using the estimated weights to reflect the target population.

   - **Weighted Brier Score ($BS_w$):** Calculated as the mean squared difference between predicted probabilities and observed outcomes, weighted by the inverse probability weights:

$$BS_w = \frac{\sum_{i=1}^{n} w_i (Y_i - \widehat{p_i})^2}{\sum_{i=1}^{n} w_i}$$

     **Variables:**
       - $Y_i = Observed\ outcome$;
       - ($\widehat{p_i}$)= Predicted probability;
       - $w_i$ = Inverse probability weight;
       - $\sum_{i=1}^{n} w_i$ = sum of all weights.

     - **Interpretation:** Measures the accuracy of probabilistic predictions. A lower score indicates better calibration and accuracy (0 = perfect, 0.25 = non-informative).

   - **Weighted Area Under the Receiver Operating Characteristic Curve ($AUC_w$):** Estimated using the weighted concordance statistic, where i indexes outcome-positive individuals and j indexes outcome-negative individuals:

$$AUC_w = \frac{\sum_{i:Y_i=1} \sum_{j:Y_j=0} w_i w_j \mathbb{1}(\widehat{p_i} > \widehat{p_j})}{\sum_{i:Y_i=1} \sum_{j:Y_j=0} w_i w_j}$$

     **Variables:**
       - $Y_i$ = Observed outcome (1 = deceased, 0 = alive)
       - ($\widehat{p_i}$)= Predicted probability;
       - $w_i$ = Inverse probability weight for patient i
       - $\mathbb{1}(\cdot)$ = Indicator function (1 if true, 0 if false)

     - **Interpretation:** Proportion of patient pairs correctly ordered by risk, weighted by IPW. Values range 0–1, where 0.5 = random chance and 1.0 = perfect discrimination.

   - **Weighted Net Benefit ($NB_W$):** Calculated at a given risk threshold ($p_t$) as the weighted True Positive count minus the penalized weighted False Positive count, normalized by the total sum of weights ($nw$):

$$NB_w = \frac{\sum_{i=1}^{n} w_i \cdot TP_i}{\sum_{i=1}^{n} w_i} - \frac{\sum_{i=1}^{n} w_i \cdot FP_i}{\sum_{i=1}^{n} w_i} \cdot \frac{p_t}{1-p_t}, \text{ where } TP_i = \mathbb{1}(\widehat{p_i} \geq p_t \wedge Y_i = 1) \text{ and } FP_i = \mathbb{1}(\widehat{p_i} \geq p_t \wedge Y_i = 0)$$

     **Variables:**
       - $p_t$= Probability threshold for intervention
       - $w_i$ = Inverse probability weight
       - $\sum_{i=1}^{n} w_i$ = sum of all weights
       - $p_t/(1 - p_t)$ = Odds at threshold (harm weighting factor)

- **Interpretation:** Net benefit represents the additional true positives per patient gained by using the model, minus the penalty for false positives weighted by threshold odds. Higher net benefit indicates better clinical utility at that threshold.

***Causal Assumptions***

The validity of using these weighted estimates to approximate the natural risk trajectory relies on three core causal inference assumptions:

- **Consistency:** We assume that the "non-intervention" state (standard ward-based care) is sufficiently well-defined and consistent across the participating hospitals, such that the potential outcome under this condition is stable.
- **Positivity (Overlap):** We assume that every patient had a non-zero probability of receiving or not receiving an intervention ($0 < P(A=1|X) < 1$). We verified this assumption by visually inspecting the distribution of propensity scores, which demonstrated substantial overlap between the intervention and non-intervention groups (see Supplementary Figure 3).
- **Exchangeability (No Unmeasured Confounding):** We assume that, conditional on the measured covariates (X), the potential outcomes are independent of the intervention assignment. To satisfy this, our propensity model included a comprehensive set of physiological parameters and deep clinical phenotyping via text embeddings to serve as proxies for a patient's medical trajectory. However, we acknowledge that unmeasured factors, such as immediate clinician intuition or real-time resource constraints, may strictly limit this assumption, rendering our results a model-based approximation of the counterfactual risk. We include this limitation in our main text's discussion section.

#### Description and Rationale for the Demographic Early Warning Score (DEWS)

***Origin and Rationale***

The Demographic Early Warning Score (DEWS) used in this study is derived directly from the International Early Warning Score (IEWS) developed and validated by Candel et al. (2023).[4] We selected this specific model framework as a primary comparator for three methodological reasons:

- **Robust Validation Metrics:** Unlike many EWS studies that rely solely on discrimination (AUC), the original IEWS was validated using a comprehensive statistical framework adhering to TRIPOD guidelines, explicitly optimizing for calibration across age groups and demonstrating clinical utility using Decision Curve Analysis (DCA). This aligns directly with the primary utility-based objective of our current study.
- **Geographical and Population Consistency:** The IEWS was developed in the Netherlands and externally validated in the Danish Multicenter Cohort (DMC). Given that our study is also conducted within the Danish healthcare system, the IEWS framework offers high ecological validity.
- **Performance in Older Populations:** The original study demonstrated that standard NEWS significantly underestimates risk in older patients (>80 years). By incorporating age and sex, the model achieved better calibration and discrimination compared to NEWS.

***Model Specification***

Our implementation of DEWS replicates the IEWS scoring system (Candel et al., 2023, Table 2) but excludes the blood pressure and temperature parameters to test our "Simplified" hypothesis. Crucially, we retained the recalibrated weights proposed by Candel et al. for the remaining parameters rather than reverting to standard NEWS weights:

- **Demographics:** Points are assigned for Age (0–7 points) and Sex (Male = 1 point) exactly as defined in the IEWS nomogram.
- **Vital Sign Recalibration:** We utilized the IEWS weights for the included vital signs (Respiratory Rate, Oxygen Saturation, Supplemental Oxygen, Pulse, Level of Consciousness). This includes specific deviations from standard NEWS to improve calibration, such as increasing the weight of altered consciousness (VPU) from 3 to 5 points and capping the weight for tachycardia (111 or higher bpm) at 2 points (see **Supplementary Table 2**).

**Modeling Approach:** DEWS was implemented as a logistic regression model using this composite score. This effectively tests whether the scoring system, combined with demographics, can compensate for the removal of time-consuming measurements (BP and Temperature).

### Supplemental Content 2 - Supplemental Tables & Figures

| Variable | Missing (N) | Missing (%) |
|---|---|---|
| Age | 0 | 0·0 |
| Sex | 0 | 0·0 |
| Previous hospitalizations | 0 | 0·0 |
| Hospital | 26 175 | 1·3 |
| Consciousness | 5 461 | 0·3 |
| Supplemental oxygen | 674 | 0·0 |
| Respiratory rate (breaths/min) | 5 494 | 0·3 |
| Pulse (beats/min) | 5 045 | 0·2 |
| Temperature (°C) | 6 472 | 0·3 |
| Oxygen saturation (%) | 5 151 | 0·2 |
| Systolic blood pressure (mm Hg) | 4 610 | 0·2 |
| Diastolic blood pressure (mm Hg) | 5 911 | 0·3 |
| Previous ICU/respiratory support | 0 | 0·0 |
| NEWS score | 8 534 | 0·4 |
| Hemoglobin (mmol/L) | 554 683 | 26·7 |
| Leukocytes ($\times 10^9$/L) | 606 741 | 29·2 |
| Platelets ($\times 10^9$/L) | 640 311 | 30·8 |
| Creatinine (µmol/L) | 546 661 | 26·3 |
| Alanine aminotransferase (U/L) | 595 636 | 28·6 |
| Lactate dehydrogenase (U/L) | 866 115 | 41·6 |
| Albumin (g/L) | 734 942 | 35·3 |
| C-reactive protein (mg/L) | 655 926 | 31·5 |
| Lactate, arterial blood (mmol/L) | 1 694 858 | 81·5 |
| Troponin T (ng/L) | 1 935 940 | 93·0 |
| Lactate, venous blood (mmol/L) | 1 923 228 | 92·4 |

**Supplementary Table 1.**
Missing data summary of variables in the final analytic cohort (N = 2 080 765) used in modelling

| Parameter | NEWS | Simplified NEWS | DEWS |
|---|---|---|---|
| **Respiratory Rate (breaths/min)** | | | |
| ≤8 | 3 | 3 | 0 |
| 9-11 | 1 | 1 | 0 |
| 12-20 | 0 | 0 | 0 |
| 21-24 | 2 | 2 | 2 |
| ≥25 | 3 | 3 | 3 |
| **Oxygen Saturation (%)** | | | |
| ≤91 | 3 | 3 | 3 |
| 92-93 | 2 | 2 | 1 |
| 94-95 | 1 | 1 | 1 |
| ≥96 | 0 | 0 | 0 |
| **Supplemental Oxygen** | | | |
| Air | 0 | 0 | 0 |
| Oxygen | 2 | 2 | 1 |
| **Systolic Blood Pressure (mmHg)** | | | |
| ≤90 | 3 | — | — |
| 91-100 | 2 | — | — |
| 101-110 | 1 | — | — |
| 111-219 | 0 | — | — |
| ≥220 | 3 | — | — |
| **Pulse Rate (beats/min)** | | | |
| ≤40 | 3 | 3 | 1 |
| 41-50 | 1 | 1 | 1 |
| 51-90 | 0 | 0 | 0 |
| 91-110 | 1 | 1 | 1 |
| 111-130 | 2 | 2 | 2 |
| ≥131 | 3 | 3 | 2 |
| **Level of Consciousness** | | | |
| Alert | 0 | 0 | 0 |
| Voice/Pain/Unresponsive | 3 | 3 | 5 |
| **Temperature (°C)** | | | |
| ≤35·0 | 3 | — | — |
| 35·1-36·0 | 1 | — | — |
| 36·1-38·0 | 0 | — | — |
| 38·1-39·0 | 1 | — | — |
| ≥39·1 | 2 | — | — |
| **Sex** | | | |
| Female | — | — | 0 |
| Male | — | — | 1 |
| **Age (years)** | | | |
| <41 | — | — | 0 |
| 41-50 | — | — | 1 |
| 51-60 | — | — | 2 |
| 61-65 | — | — | 3 |
| 66-75 | — | — | 4 |
| 76-80 | — | — | 5 |
| 81-90 | — | — | 6 |
| ≥91 | — | — | 7 |
| **Total Score Range** | | | |
| 0-20 | 0-20 | 0-14 | 0-22 |
| — indicates parameter not included in scoring system. All scoring systems used the first recorded vital sign measurement per hospitalization. | | | |

**Supplementary Table 2.**
Analytical description of the scoring systems used in the current study

| Parameter | Parameter Name | Value |
|---|---|---|
| `tree_depth` | Maximum Tree Depth | 6 |
| `trees` | Number of Trees | 100 |
| `learn_rate` | Learning Rate | 0·3 |
| `mtry` | Predictors per Split | 30 |
| `min_n` | Minimal Node Size | 1 |
| `loss_reduction` | Minimum Loss Reduction | 0 |
| `sample_size` | Proportion of Data Sampled | 1 |
| Default values from the tidymodels engine were used, with the exception of 'Number of Trees' and 'Predictors per Split'. | | |

**Supplementary Table 3.**
XGB-EWS hyperparameter configuration

| NEWS Clinical Risk Group | Response | Count (N) | Observed Weighted 24-Hour Mortality Risk | Mean Predicted Risk (NEWS) | Corresponding Decision Curve Analysis Threshold | XGB-EWS Risk Range (Predicted Probability) |
|---|---|---|---|---|---|---|
| **Low** (0–4) | Ward-based response (Min 4–12 hourly monitoring) | 1 879 031 (90·6%) | 0·06% (0·000644) | 0·08% (0·0008) | < 0·0022 | Risk < 0·22% |
| **Low-Medium** (Score of 3 in any individual parameter) | Urgent ward-based response (Min 1 hourly monitoring) | 60 892 (2·9%) | 0·36% (0·00363) | 0·25% (0·0025) | 0·0044 | Risk in [0·22%, 0·88%) |
| **Medium** (5–6) | Key threshold for urgent response (Min 1 hourly monitoring) | 90 686 (4·2%) | 0·93% (0·00930) | 0·72% (0·0072) | 0·0088 | Risk in [0·88%, 4·41%) |
| **High** (≥7) | Urgent or emergency response (Continuous monitoring) | 50 156 (2·3%) | 4·89% (0·0489) | 4·95% (0·0495) | 0·0441 | Risk ≥ 4·41% |

**Supplementary Table 4.**
Alignment of National Early Warning Score (NEWS) Clinical Risk Categories with Observed 24-Hour Mortality and Decision Curve Analysis Thresholds. The table maps the standard NEWS clinical escalation categories to the observed weighted and predicted 24-hour mortality risk within the study cohort. Count (N) represents the number of hospital encounters along with the weighted percentages in each risk group. Observed Weighted Risk reflects the mortality prevalence after correcting for intervention bias. Corresponding Decision Curve Analysis Threshold identifies the specific risk threshold used in the net benefit analysis that most closely approximates the observed risk for that clinical group. Risk ranges of XGB-EWS corresponding to NEWS clinical risk groups are also presented.

| | Net Benefit per 10 000 Patients | | | | | | Net Benefit Difference per 10 000 Patients | | | |
|---|---|---|---|---|---|---|---|---|---|---|
| Risk Threshold | Treat All | Treat None | NEWS | Simplified NEWS | DEWS | XGB-EWS | NEWS - XGB-EWS | NEWS - Simplified NEWS | DEWS - XGB-EWS | NEWS - DEWS |
| **18-65 (N: 1 053 140, 50·9%)** | | | | | | | | | | |
| 0·0005 | 0·0 | 0·0 | 2·2 | 1·4 | 3·4 | 3·7 | -1·5 | 0·8 | -0·3 | -1·2 |
| 0·0010 | -5·0 | 0·0 | 1·9 | 2·4 | 2·9 | 3·4 | -1·5 | -0·5 | -0·5 | -1·0 |
| 0·0020 | -15·1 | 0·0 | 2·2 | 1·9 | 2·4 | 2·9 | -0·7 | 0·3 | -0·5 | -0·2 |
| 0·0040 | -35·4 | 0·0 | 1·8 | 1·4 | 1·7 | 2·5 | -0·7 | 0·4 | -0·8 | 0·1 |
| 0·0050 | -45·5 | 0·0 | 1·4 | 1·2 | 1·6 | 2·3 | -0·9 | 0·2 | -0·7 | -0·2 |
| 0·0101 | -96·6 | 0·0 | 1·2 | 0·8 | 1·0 | 1·8 | -0·6 | 0·4 | -0·8 | 0·2 |
| 0·0151 | -148·2 | 0·0 | 0·7 | 0·2 | 0·9 | 1·6 | -0·9 | 0·5 | -0·7 | -0·2 |
| **66-80 (N: 629 221, 30·6%)** | | | | | | | | | | |
| 0·0027 | 0·0 | 0·0 | 16·9 | 15·7 | 15·5 | 17·5 | -0·6 | 1·2 | -2·0 | 1·4 |
| 0·0053 | -26·8 | 0·0 | 13·6 | 12·2 | 12·7 | 14·7 | -1·1 | 1·4 | -2·0 | 0·9 |
| 0·0107 | -80·7 | 0·0 | 10·3 | 9·0 | 8·9 | 11·8 | -1·5 | 1·3 | -2·9 | 1·4 |
| 0·0213 | -190·5 | 0·0 | 7·4 | 6·2 | 6·3 | 9·1 | -1·7 | 1·2 | -2·8 | 1·1 |
| 0·0266 | -246·2 | 0·0 | 6·4 | 5·1 | 5·2 | 8·2 | -1·8 | 1·3 | -3·0 | 1·2 |
| 0·0533 | -534·4 | 0·0 | 3·9 | 3·1 | 3·6 | 5·5 | -1·6 | 0·8 | -1·9 | 0·3 |
| 0·0799 | -839·3 | 0·0 | 3·0 | 1·7 | 2·4 | 4·3 | -1·3 | 1·3 | -1·9 | 0·6 |
| **80+ (N: 398 404, 18·5%)** | | | | | | | | | | |
| 0·0061 | 0·0 | 0·0 | 33·7 | 32·1 | 34·7 | 38·1 | -4·4 | 1·6 | -3·4 | -1·0 |
| 0·0122 | -61·9 | 0·0 | 26·9 | 24·0 | 27·2 | 31·0 | -4·1 | 2·9 | -3·8 | -0·3 |
| 0·0245 | -188·2 | 0·0 | 20·1 | 16·8 | 19·3 | 24·4 | -4·3 | 3·3 | -5·1 | 0·8 |
| 0·0489 | -450·3 | 0·0 | 11·8 | 8·4 | 11·4 | 18·2 | -6·4 | 3·4 | -6·8 | 0·4 |
| 0·0612 | -586·6 | 0·0 | 10·7 | 7·6 | 9·5 | 16·1 | -5·4 | 3·1 | -6·6 | 1·2 |
| 0·1224 | -1 324·6 | 0·0 | 6·7 | 3·8 | 4·1 | 9·7 | -3·0 | 2·9 | -5·6 | 2·6 |
| 0·1836 | -2 173·3 | 0·0 | 4·0 | 2·4 | 1·6 | 6·4 | -2·4 | 1·6 | -4·8 | 2·4 |
| Positive values of net benefit difference indicate the first model has higher net benefit; negative values indicate the comparator is better. Risk thresholds are chosen as multiples of the age-specific prevalence. | | | | | | | | | | |

**Supplementary Table 5.**
Net Benefit comparison of different strategies and models across age groups at specified risk thresholds.

| | Net Benefit per 10 000 Patients | | | | | | Net Benefit Difference per 10 000 Patients | | | |
|---|---|---|---|---|---|---|---|---|---|---|
| Risk Threshold | Treat All | Treat None | NEWS | Simplified NEWS | DEWS | XGB-EWS | NEWS - XGB-EWS | NEWS - Simplified NEWS | DEWS - XGB-EWS | NEWS - DEWS |
| **Female (N: 1 043 017, 50·4%)** | | | | | | | | | | |
| 0·0020 | 0·0 | 0·0 | 13·2 | 12·8 | 13·1 | 14·3 | -1·1 | 0·4 | -1·2 | 0·1 |
| 0·0039 | -19·7 | 0·0 | 11·3 | 10·6 | 11·1 | 12·3 | -1·0 | 0·7 | -1·2 | 0·2 |
| 0·0078 | -59·3 | 0·0 | 9·3 | 8·3 | 8·7 | 10·3 | -1·0 | 1·0 | -1·6 | 0·6 |
| 0·0157 | -139·4 | 0·0 | 7·0 | 5·2 | 5·8 | 8·0 | -1·0 | 1·8 | -2·2 | 1·2 |
| 0·0196 | -180·0 | 0·0 | 5·9 | 4·8 | 5·4 | 7·4 | -1·5 | 1·1 | -2·0 | 0·5 |
| 0·0392 | -387·7 | 0·0 | 3·9 | 2·7 | 3·6 | 5·4 | -1·5 | 1·2 | -1·8 | 0·3 |
| 0·0588 | -604·1 | 0·0 | 2·7 | 1·9 | 2·6 | 4·4 | -1·7 | 0·8 | -1·8 | 0·1 |
| **Male (N: 1 037 748, 49·6%)** | | | | | | | | | | |
| 0·0025 | 0·0 | 0·0 | 16·0 | 15·4 | 15·7 | 17·2 | -1·2 | 0·6 | -1·5 | 0·3 |
| 0·0049 | -24·6 | 0·0 | 13·3 | 12·1 | 13·2 | 14·8 | -1·5 | 1·2 | -1·6 | 0·1 |
| 0·0098 | -74·3 | 0·0 | 10·0 | 8·9 | 9·8 | 12·1 | -2·1 | 1·1 | -2·3 | 0·2 |
| 0·0196 | -175·1 | 0·0 | 7·4 | 6·3 | 7·3 | 9·6 | -2·2 | 1·1 | -2·3 | 0·1 |
| 0·0245 | -226·2 | 0·0 | 6·7 | 5·6 | 6·5 | 8·8 | -2·1 | 1·1 | -2·3 | 0·2 |
| 0·0490 | -489·9 | 0·0 | 4·0 | 2·9 | 4·0 | 6·3 | -2·3 | 1·1 | -2·3 | 0·0 |
| 0·0736 | -767·5 | 0·0 | 3·0 | 2·1 | 2·8 | 4·9 | -1·9 | 0·9 | -2·1 | 0·2 |
| Positive values of net benefit difference indicate the first model has higher net benefit; negative values indicate the comparator is better. Risk thresholds are chosen as multiples of the sex-specific prevalence. | | | | | | | | | | |

**Supplementary Table 6.**
Net benefit comparison of different strategies and models across sex groups at specified risk thresholds.

| | Net Benefit per 10 000 Patients | | | | | | Net Benefit Difference per 10 000 Patients | | | |
|---|---|---|---|---|---|---|---|---|---|---|
| **Risk Threshold** | **Treat All** | **Treat None** | **NEWS** | **Simplified NEWS** | **DEWS** | **XGB-EWS** | **NEWS - XGB-EWS** | **NEWS - Simplified NEWS** | **DEWS - XGB-EWS** | **NEWS - DEWS** |
| **Cardiopulmonary (N: 151 050, 7·4%)** | | | | | | | | | | |
| 0·0015 | 0·0 | 0·0 | 8·8 | 8·4 | 8·2 | 10·0 | -1·2 | 0·4 | -1·8 | 0·6 |
| 0·0030 | -14·9 | 0·0 | 6·4 | 5·9 | 5·9 | 8·0 | -1·6 | 0·5 | -2·1 | 0·5 |
| 0·0059 | -44·7 | 0·0 | 4·7 | 3·7 | 4·3 | 6·4 | -1·7 | 1·0 | -2·1 | 0·4 |
| 0·0119 | -105·0 | 0·0 | 3·0 | 1·8 | 3·1 | 4·6 | -1·6 | 1·2 | -1·5 | -0·1 |
| 0·0148 | -135·4 | 0·0 | 2·1 | 0·7 | 2·2 | 4·2 | -2·1 | 1·4 | -2·0 | -0·1 |
| 0·0297 | -290·3 | 0·0 | 1·0 | 0·1 | 1·7 | 2·6 | -1·6 | 0·9 | -0·9 | -0·7 |
| 0·0445 | -450·0 | 0·0 | -0·5 | -1·6 | 1·0 | 1·9 | -2·4 | 1·1 | -0·9 | -1·5 |
| **Critical Care (N: 64 262, 3·2%)** | | | | | | | | | | |
| 0·0006 | 0·0 | 0·0 | 1·8 | 2·5 | 3·3 | 4·5 | -2·7 | -0·7 | -1·2 | -1·5 |
| 0·0012 | -6·1 | 0·0 | 2·9 | 1·5 | 2·3 | 4·1 | -1·2 | 1·4 | -1·8 | 0·6 |
| 0·0024 | -18·2 | 0·0 | 2·4 | 1·7 | 2·9 | 3·8 | -1·4 | 0·7 | -0·9 | -0·5 |
| 0·0048 | -42·6 | 0·0 | 1·9 | 1·5 | 2·2 | 3·2 | -1·3 | 0·4 | -1·0 | -0·3 |
| 0·0061 | -54·9 | 0·0 | 2·9 | 2·2 | 2·4 | 2·9 | 0·0 | 0·7 | -0·5 | 0·5 |
| 0·0121 | -116·6 | 0·0 | 1·9 | 1·8 | 1·6 | 2·2 | -0·3 | 0·1 | -0·6 | 0·3 |
| 0·0182 | -179·1 | 0·0 | 1·3 | 0·9 | 1·1 | 1·8 | -0·5 | 0·4 | -0·7 | 0·2 |
| **Gastroenterology (N: 20 518, 1·0%)** | | | | | | | | | | |
| 0·0013 | 0·0 | 0·0 | 7·7 | 7·7 | 8·1 | 9·9 | -2·2 | 0·0 | -1·8 | -0·4 |
| 0·0026 | -13·3 | 0·0 | 7·2 | 5·7 | 5·4 | 8·6 | -1·4 | 1·5 | -3·2 | 1·8 |
| 0·0053 | -39·9 | 0·0 | 5·6 | 3·9 | 4·2 | 6·4 | -0·8 | 1·7 | -2·2 | 1·4 |
| 0·0106 | -93·6 | 0·0 | 3·8 | 2·0 | 1·3 | 5·3 | -1·5 | 1·8 | -4·0 | 2·5 |
| 0·0132 | -120·6 | 0·0 | 3·1 | 1·3 | 0·7 | 4·4 | -1·3 | 1·8 | -3·7 | 2·4 |
| 0·0264 | -258·1 | 0·0 | 0·3 | 0·1 | 0·5 | 2·0 | -1·7 | 0·2 | -1·5 | -0·2 |
| 0·0397 | -399·3 | 0·0 | -0·5 | -0·7 | -0·3 | 1·6 | -2·1 | 0·2 | -1·9 | -0·2 |
| **Internal & Acute Medicine (N: 1 382 512, 67·5%)** | | | | | | | | | | |
| 0·0027 | 0·0 | 0·0 | 18·0 | 17·3 | 17·8 | 19·1 | -1·1 | 0·7 | -1·3 | 0·2 |
| 0·0054 | -27·0 | 0·0 | 15·2 | 13·6 | 15·1 | 16·4 | -1·2 | 1·6 | -1·3 | 0·1 |
| 0·0108 | -81·6 | 0·0 | 11·6 | 10·2 | 11·2 | 13·5 | -1·9 | 1·4 | -2·3 | 0·4 |
| 0·0215 | -192·4 | 0·0 | 8·7 | 7·3 | 8·2 | 10·7 | -2·0 | 1·4 | -2·5 | 0·5 |
| 0·0269 | -248·7 | 0·0 | 8·0 | 6·2 | 7·0 | 9·7 | -1·7 | 1·8 | -2·7 | 1·0 |
| 0·0538 | -540·0 | 0·0 | 4·9 | 3·6 | 4·6 | 7·0 | -2·1 | 1·3 | -2·4 | 0·3 |
| 0·0807 | -848·3 | 0·0 | 3·7 | 2·4 | 3·3 | 5·6 | -1·9 | 1·3 | -2·3 | 0·4 |
| **Nephrology (N: 15 055, 0·7%)** | | | | | | | | | | |
| 0·0013 | 0·0 | 0·0 | 8·8 | 8·1 | 9·1 | 7·9 | 0·9 | 0·7 | 1·2 | -0·3 |
| 0·0027 | -13·3 | 0·0 | 7·0 | 7·1 | 7·1 | 5·7 | 1·3 | -0·1 | 1·4 | -0·1 |
| 0·0053 | -40·0 | 0·0 | 4·5 | 3·2 | 5·7 | 4·7 | -0·2 | 1·3 | 1·0 | -1·2 |
| 0·0106 | -93·8 | 0·0 | 2·5 | 2·6 | 3·8 | 3·2 | -0·7 | -0·1 | 0·6 | -1·3 |
| 0·0133 | -121·0 | 0·0 | 1·8 | 2·0 | 3·3 | 3·3 | -1·5 | -0·2 | 0·0 | -1·5 |
| 0·0265 | -258·9 | 0·0 | 0·5 | -0·5 | 0·9 | 2·9 | -2·4 | 1·0 | -2·0 | -0·4 |
| 0·0398 | -400·6 | 0·0 | 0·3 | -0·5 | 0·1 | 1·7 | -1·4 | 0·8 | -1·6 | 0·2 |
| **Neurology (N: 54 715, 2·8%)** | | | | | | | | | | |
| 0·0018 | 0·0 | 0·0 | 12·4 | 11·8 | 11·0 | 11·8 | 0·6 | 0·6 | -0·8 | 1·4 |
| 0·0036 | -17·8 | 0·0 | 9·1 | 8·8 | 9·6 | 9·6 | -0·5 | 0·3 | 0·0 | -0·5 |
| 0·0071 | -53·7 | 0·0 | 6·2 | 7·5 | 7·7 | 8·3 | -2·1 | -1·3 | -0·6 | -1·5 |
| 0·0142 | -126·2 | 0·0 | 4·2 | 3·3 | 6·7 | 6·1 | -1·9 | 0·9 | 0·6 | -2·5 |
| 0·0178 | -162·8 | 0·0 | 2·8 | 3·0 | 4·6 | 5·7 | -2·9 | -0·2 | -1·1 | -1·8 |
| 0·0355 | -350·1 | 0·0 | 1·2 | 0·9 | 1·8 | 3·7 | -2·5 | 0·3 | -1·9 | -0·6 |
| 0·0533 | -544·4 | 0·0 | 0·8 | 0·6 | 0·5 | 2·6 | -1·8 | 0·2 | -2·1 | 0·3 |
| **Oncology & Hematology (N: 48 048, 2·4%)** | | | | | | | | | | |
| 0·0020 | 0·0 | 0·0 | 13·0 | 12·1 | 10·1 | 12·7 | 0·3 | 0·9 | -2·6 | 2·9 |
| 0·0041 | -20·4 | 0·0 | 10·9 | 9·9 | 7·5 | 10·2 | 0·7 | 1·0 | -2·7 | 3·4 |
| 0·0081 | -61·4 | 0·0 | 8·5 | 7·5 | 5·5 | 8·1 | 0·4 | 1·0 | -2·6 | 3·0 |
| 0·0162 | -144·5 | 0·0 | 6·2 | 3·7 | 2·9 | 6·1 | 0·1 | 2·5 | -3·2 | 3·3 |
| 0·0203 | -186·6 | 0·0 | 4·3 | 3·2 | 2·7 | 5·8 | -1·5 | 1·1 | -3·1 | 1·6 |

| | Net Benefit per 10 000 Patients | | | | | | Net Benefit Difference per 10 000 Patients | | | |
|---|---|---|---|---|---|---|---|---|---|---|
| Risk Threshold | Treat All | Treat None | NEWS | Simplified NEWS | DEWS | XGB-EWS | NEWS - XGB-EWS | NEWS - Simplified NEWS | DEWS - XGB-EWS | NEWS - DEWS |
| 0·0406 | -402·3 | 0·0 | 1·7 | 2·0 | 1·5 | 3·6 | -1·9 | -0·3 | -2·1 | 0·2 |
| 0·0609 | -627·3 | 0·0 | 1·5 | 0·9 | 1·0 | 3·2 | -1·7 | 0·6 | -2·2 | 0·5 |
| **Surgical (N: 303 709, 15·0%)** | | | | | | | | | | |
| 0·0011 | 0·0 | 0·0 | 7·0 | 7·1 | 6·7 | 8·2 | -1·2 | -0·1 | -1·5 | 0·3 |
| 0·0022 | -11·0 | 0·0 | 6·4 | 6·0 | 5·9 | 7·4 | -1·0 | 0·4 | -1·5 | 0·5 |
| 0·0044 | -33·2 | 0·0 | 5·2 | 4·4 | 4·9 | 6·3 | -1·1 | 0·8 | -1·4 | 0·3 |
| 0·0088 | -77·9 | 0·0 | 4·0 | 3·3 | 3·5 | 4·9 | -0·9 | 0·7 | -1·4 | 0·5 |
| 0·0110 | -100·3 | 0·0 | 3·3 | 2·7 | 2·9 | 4·3 | -1·0 | 0·6 | -1·4 | 0·4 |
| 0·0221 | -214·2 | 0·0 | 1·9 | 1·7 | 1·9 | 3·2 | -1·3 | 0·2 | -1·3 | 0·0 |
| 0·0331 | -330·7 | 0·0 | 1·2 | 1·1 | 1·2 | 2·5 | -1·3 | 0·1 | -1·3 | 0·0 |
| Positive values of net benefit difference indicate the first model has higher net benefit; negative values indicate the comparator is better. Risk thresholds are chosen as multiples of the clinical specialty-specific prevalence. | | | | | | | | | | |

**Supplementary Table 7.**
Net Benefit comparison of different strategies and models across clinical specialties at specified risk thresholds.

| | Net Benefit per 10 000 Patients | | | | | | Net Benefit Difference per 10 000 Patients | | | |
|---|---|---|---|---|---|---|---|---|---|---|
| Risk Threshold | Treat All | Treat None | NEWS | Simplified NEWS | DEWS | XGB-EWS | NEWS - XGB-EWS | NEWS - Simplified NEWS | DEWS - XGB-EWS | NEWS - DEWS |
| **Certain infectious and parasitic diseases [A00-B99] (N: 81 079, 4·8%)** | | | | | | | | | | |
| 0·0039 | 0·0 | 0·0 | 25·0 | 24·5 | 25·4 | 26·9 | -1·9 | 0·5 | -1·5 | -0·4 |
| 0·0078 | -39·3 | 0·0 | 21·8 | 20·9 | 20·7 | 22·9 | -1·1 | 0·9 | -2·2 | 1·1 |
| 0·0156 | -118·9 | 0·0 | 17·8 | 14·9 | 17·0 | 19·7 | -1·9 | 2·9 | -2·7 | 0·8 |
| 0·0312 | -281·8 | 0·0 | 13·8 | 11·2 | 12·3 | 15·3 | -1·5 | 2·6 | -3·0 | 1·5 |
| 0·0390 | -365·3 | 0·0 | 12·3 | 9·9 | 10·9 | 13·9 | -1·6 | 2·4 | -3·0 | 1·4 |
| 0·0780 | -803·8 | 0·0 | 6·4 | 4·2 | 4·9 | 9·1 | -2·7 | 2·2 | -4·2 | 1·5 |
| 0·1170 | -1 281·1 | 0·0 | 4·5 | 2·8 | 2·7 | 7·3 | -2·8 | 1·7 | -4·6 | 1·8 |
| **Diseases of the blood and blood-forming organs and certain disorders involving the immune mechanism [D50-D89] (N: 17 373, 1·0%)** | | | | | | | | | | |
| 0·0035 | 0·0 | 0·0 | 22·9 | 19·8 | 19·2 | 23·5 | -0·6 | 3·1 | -4·3 | 3·7 |
| 0·0069 | -35·0 | 0·0 | 19·3 | 13·9 | 14·0 | 21·0 | -1·7 | 5·4 | -7·0 | 5·3 |
| 0·0139 | -105·7 | 0·0 | 14·0 | 12·6 | 11·0 | 16·4 | -2·4 | 1·4 | -5·4 | 3·0 |
| 0·0278 | -250·1 | 0·0 | 8·3 | 4·2 | 8·6 | 13·7 | -5·4 | 4·1 | -5·1 | -0·3 |
| 0·0347 | -323·9 | 0·0 | 7·4 | 4·0 | 8·2 | 10·5 | -3·1 | 3·4 | -2·3 | -0·8 |
| 0·0695 | -709·4 | 0·0 | 3·4 | 3·5 | 4·3 | 5·2 | -1·8 | -0·1 | -0·9 | -0·9 |
| 0·1042 | -1 124·7 | 0·0 | 3·4 | 2·6 | 3·7 | 2·9 | 0·5 | 0·8 | 0·8 | -0·3 |
| **Diseases of the circulatory system [I00-I99] (N: 169 102, 10·4%)** | | | | | | | | | | |
| 0·0038 | 0·0 | 0·0 | 22·0 | 20·5 | 22·6 | 25·1 | -3·1 | 1·5 | -2·5 | -0·6 |
| 0·0076 | -38·4 | 0·0 | 18·2 | 16·7 | 18·1 | 21·8 | -3·6 | 1·5 | -3·7 | 0·1 |
| 0·0152 | -116·1 | 0·0 | 14·1 | 11·0 | 14·8 | 17·4 | -3·3 | 3·1 | -2·6 | -0·7 |
| 0·0305 | -275·2 | 0·0 | 8·2 | 6·2 | 8·6 | 13·5 | -5·3 | 2·0 | -4·9 | -0·4 |
| 0·0381 | -356·7 | 0·0 | 7·7 | 5·6 | 8·0 | 11·9 | -4·2 | 2·1 | -3·9 | -0·3 |
| 0·0762 | -784·1 | 0·0 | 4·9 | 2·9 | 4·3 | 8·2 | -3·3 | 2·0 | -3·9 | 0·6 |
| 0·1144 | -1 248·2 | 0·0 | 3·6 | 1·6 | 2·5 | 6·7 | -3·1 | 2·0 | -4·2 | 1·1 |
| **Diseases of the digestive system [K00-K93] (N: 108 687, 6·6%)** | | | | | | | | | | |
| 0·0020 | 0·0 | 0·0 | 12·1 | 11·5 | 12·1 | 13·7 | -1·6 | 0·6 | -1·6 | 0·0 |
| 0·0040 | -20·0 | 0·0 | 10·3 | 9·7 | 9·9 | 11·0 | -0·7 | 0·6 | -1·1 | 0·4 |
| 0·0080 | -60·1 | 0·0 | 8·3 | 7·2 | 7·2 | 8·9 | -0·6 | 1·1 | -1·7 | 1·1 |
| 0·0159 | -141·4 | 0·0 | 6·8 | 4·1 | 4·3 | 6·4 | 0·4 | 2·7 | -2·1 | 2·5 |
| 0·0199 | -182·5 | 0·0 | 4·9 | 3·8 | 3·8 | 6·1 | -1·2 | 1·1 | -2·3 | 1·1 |
| 0·0398 | -393·3 | 0·0 | 3·0 | 1·6 | 2·2 | 4·2 | -1·2 | 1·4 | -2·0 | 0·8 |
| 0·0596 | -613·1 | 0·0 | 2·0 | 0·9 | 1·0 | 3·0 | -1·0 | 1·1 | -2·0 | 1·0 |
| **Diseases of the ear and mastoid process [H60-H95] (N: 7 325, 0·5%)** | | | | | | | | | | |
| 0·0001 | 0·0 | 0·0 | 0·0 | 0·0 | 0·3 | 0·7 | -0·7 | 0·0 | -0·4 | -0·3 |
| 0·0002 | -1·2 | 0·0 | -1·2 | -1·2 | -0·4 | 0·6 | -1·8 | 0·0 | -1·0 | -0·8 |
| 0·0005 | -3·7 | 0·0 | -0·9 | -2·2 | -0·7 | 0·5 | -1·4 | 1·3 | -1·2 | -0·2 |
| 0·0010 | -8·7 | 0·0 | -0·5 | 0·1 | -1·2 | 0·6 | -1·1 | -0·6 | -1·8 | 0·7 |
| 0·0012 | -11·2 | 0·0 | 0·3 | -0·2 | -1·1 | 0·6 | -0·3 | 0·5 | -1·7 | 1·4 |
| 0·0025 | -23·8 | 0·0 | 0·5 | 0·1 | 0·1 | 0·6 | -0·1 | 0·4 | -0·5 | 0·4 |
| 0·0037 | -36·3 | 0·0 | 0·7 | 0·6 | 0·6 | 0·7 | 0·0 | 0·1 | -0·1 | 0·1 |
| **Diseases of the eye and adnexa [H00-H59] (N: 4 819, 0·3%)** | | | | | | | | | | |
| 0·0006 | 0·0 | 0·0 | 3·5 | 4·1 | 3·4 | 5·0 | -1·5 | -0·6 | -1·6 | 0·1 |
| 0·0012 | -6·0 | 0·0 | 4·9 | 4·4 | 2·7 | 4·9 | 0·0 | 0·5 | -2·2 | 2·2 |
| 0·0024 | -18·0 | 0·0 | 4·9 | 4·5 | 4·5 | 4·8 | 0·1 | 0·4 | -0·3 | 0·4 |
| 0·0048 | -42·1 | 0·0 | 4·8 | 4·7 | 4·7 | 4·7 | 0·1 | 0·1 | 0·0 | 0·1 |
| 0·0060 | -54·2 | 0·0 | 3·3 | 5·1 | 5·1 | 1·0 | 2·3 | -1·8 | 4·1 | -1·8 |
| 0·0120 | -115·1 | 0·0 | 3·1 | 3·0 | 1·1 | 1·0 | 2·1 | 0·1 | 0·1 | 2·0 |
| 0·0180 | -176·8 | 0·0 | 1·3 | 1·2 | 1·1 | 0·9 | 0·4 | 0·1 | 0·2 | 0·2 |
| **Diseases of the genitourinary system [N00-N99] (N: 92 927, 5·7%)** | | | | | | | | | | |
| 0·0015 | 0·0 | 0·0 | 9·8 | 8·9 | 8·9 | 9·8 | 0·0 | 0·9 | -0·9 | 0·9 |
| 0·0031 | -15·4 | 0·0 | 7·9 | 6·8 | 7·3 | 8·8 | -0·9 | 1·1 | -1·5 | 0·6 |
| 0·0061 | -46·4 | 0·0 | 5·9 | 5·4 | 5·0 | 7·4 | -1·5 | 0·5 | -2·4 | 0·9 |
| 0·0123 | -108·9 | 0·0 | 4·5 | 3·5 | 2·9 | 5·5 | -1·0 | 1·0 | -2·6 | 1·6 |
| 0·0154 | -140·5 | 0·0 | 3·9 | 2·6 | 2·3 | 5·1 | -1·2 | 1·3 | -2·8 | 1·6 |

| | Net Benefit per 10 000 Patients | | | | | | Net Benefit Difference per 10 000 Patients | | | |
|---|---|---|---|---|---|---|---|---|---|---|
| Risk Threshold | Treat All | Treat None | NEWS | Simplified NEWS | DEWS | XGB-EWS | NEWS - XGB-EWS | NEWS - Simplified NEWS | DEWS - XGB-EWS | NEWS - DEWS |
| 0·0307 | -301·3 | 0·0 | 2·6 | 1·1 | 1·6 | 3·9 | -1·3 | 1·5 | -2·3 | 1·0 |
| 0·0461 | -467·3 | 0·0 | 1·7 | 0·3 | 1·1 | 2·9 | -1·2 | 1·4 | -1·8 | 0·6 |
| **Diseases of the musculoskeletal system and connective tissue [M00-M99] (N: 78 050, 4·8%)** | | | | | | | | | | |
| 0·0009 | 0·0 | 0·0 | 6·4 | 4·8 | 5·6 | 7·2 | -0·8 | 1·6 | -1·6 | 0·8 |
| 0·0017 | -8·7 | 0·0 | 6·2 | 5·9 | 5·1 | 6·5 | -0·3 | 0·3 | -1·4 | 1·1 |
| 0·0035 | -26·2 | 0·0 | 4·9 | 4·6 | 4·5 | 5·8 | -0·9 | 0·3 | -1·3 | 0·4 |
| 0·0070 | -61·3 | 0·0 | 4·0 | 3·6 | 3·5 | 4·3 | -0·3 | 0·4 | -0·8 | 0·5 |
| 0·0087 | -79·0 | 0·0 | 3·7 | 2·8 | 3·2 | 3·9 | -0·2 | 0·9 | -0·7 | 0·5 |
| 0·0174 | -168·2 | 0·0 | 2·1 | 1·6 | 1·8 | 2·8 | -0·7 | 0·5 | -1·0 | 0·3 |
| 0·0261 | -259·0 | 0·0 | 1·7 | 1·2 | 1·3 | 1·8 | -0·1 | 0·5 | -0·5 | 0·4 |
| **Diseases of the nervous system [G00-G99] (N: 34 196, 2·1%)** | | | | | | | | | | |
| 0·0011 | 0·0 | 0·0 | 7·3 | 7·3 | 6·1 | 7·5 | -0·2 | 0·0 | -1·4 | 1·2 |
| 0·0021 | -10·5 | 0·0 | 6·2 | 6·4 | 5·7 | 6·0 | 0·2 | -0·2 | -0·3 | 0·5 |
| 0·0042 | -31·7 | 0·0 | 4·8 | 4·7 | 4·8 | 5·0 | -0·2 | 0·1 | -0·2 | 0·0 |
| 0·0084 | -74·2 | 0·0 | 3·3 | 3·2 | 2·4 | 4·3 | -1·0 | 0·1 | -1·9 | 0·9 |
| 0·0105 | -95·6 | 0·0 | 2·9 | 2·8 | 2·3 | 3·1 | -0·2 | 0·1 | -0·8 | 0·6 |
| 0·0210 | -204·0 | 0·0 | 0·5 | 0·7 | 0·5 | 3·1 | -2·6 | -0·2 | -2·6 | 0·0 |
| 0·0315 | -314·8 | 0·0 | 0·5 | 0·1 | 0·6 | 2·2 | -1·7 | 0·4 | -1·6 | -0·1 |
| **Diseases of the respiratory system [J00-J99] (N: 110 252, 6·6%)** | | | | | | | | | | |
| 0·0040 | 0·0 | 0·0 | 23·6 | 22·3 | 24·5 | 26·2 | -2·6 | 1·3 | -1·7 | -0·9 |
| 0·0079 | -40·0 | 0·0 | 17·3 | 15·9 | 18·1 | 21·1 | -3·8 | 1·4 | -3·0 | -0·8 |
| 0·0159 | -120·9 | 0·0 | 11·4 | 11·7 | 14·0 | 15·7 | -4·3 | -0·3 | -1·7 | -2·6 |
| 0·0317 | -286·8 | 0·0 | 8·6 | 7·0 | 9·6 | 10·5 | -1·9 | 1·6 | -0·9 | -1·0 |
| 0·0397 | -371·7 | 0·0 | 6·3 | 4·1 | 7·2 | 9·3 | -3·0 | 2·2 | -2·1 | -0·9 |
| 0·0793 | -818·6 | 0·0 | 3·6 | 0·8 | 5·2 | 5·1 | -1·5 | 2·8 | 0·1 | -1·6 |
| 0·1190 | -1 305·7 | 0·0 | 1·0 | 0·3 | 2·4 | 2·8 | -1·8 | 0·7 | -0·4 | -1·4 |
| **Diseases of the skin and subcutaneous tissue [L00-L99] (N: 16 491, 1·0%)** | | | | | | | | | | |
| 0·0006 | 0·0 | 0·0 | 2·4 | 3·1 | 3·4 | 4·4 | -2·0 | -0·7 | -1·0 | -1·0 |
| 0·0012 | -6·0 | 0·0 | 4·0 | 3·5 | 2·8 | 4·3 | -0·3 | 0·5 | -1·5 | 1·2 |
| 0·0024 | -18·1 | 0·0 | 4·0 | 3·0 | 3·2 | 3·6 | 0·4 | 1·0 | -0·4 | 0·8 |
| 0·0048 | -42·4 | 0·0 | 3·8 | 3·0 | 2·5 | 3·5 | 0·3 | 0·8 | -1·0 | 1·3 |
| 0·0060 | -54·5 | 0·0 | 3·7 | 3·5 | 3·0 | 3·5 | 0·2 | 0·2 | -0·5 | 0·7 |
| 0·0120 | -115·8 | 0·0 | 3·5 | 3·4 | 2·8 | 2·8 | 0·7 | 0·1 | 0·0 | 0·7 |
| 0·0181 | -177·8 | 0·0 | 3·1 | 2·3 | 3·0 | 1·6 | 1·5 | 0·8 | 1·4 | 0·1 |
| **Endocrine, nutritional and metabolic diseases [E00-E90] (N: 45 626, 2·8%)** | | | | | | | | | | |
| 0·0027 | 0·0 | 0·0 | 14·4 | 14·7 | 15·4 | 17·2 | -2·8 | -0·3 | -1·8 | -1·0 |
| 0·0053 | -26·9 | 0·0 | 12·2 | 8·9 | 11·9 | 13·2 | -1·0 | 3·3 | -1·3 | 0·3 |
| 0·0107 | -81·1 | 0·0 | 8·0 | 6·2 | 6·8 | 11·6 | -3·6 | 1·8 | -4·8 | 1·2 |
| 0·0214 | -191·3 | 0·0 | 6·1 | 4·1 | 3·7 | 7·8 | -1·7 | 2·0 | -4·1 | 2·4 |
| 0·0267 | -247·3 | 0·0 | 5·3 | 2·9 | 3·3 | 6·8 | -1·5 | 2·4 | -3·5 | 2·0 |
| 0·0535 | -536·8 | 0·0 | 2·7 | 1·8 | 1·5 | 3·0 | -0·3 | 0·9 | -1·5 | 1·2 |
| 0·0802 | -843·1 | 0·0 | 1·4 | 1·1 | 0·8 | 2·2 | -0·8 | 0·3 | -1·4 | 0·6 |
| **Factors influencing health status and contact with health services [Z00-Z99] (N: 180 695, 11·0%)** | | | | | | | | | | |
| 0·0020 | 0·0 | 0·0 | 12·8 | 13·0 | 13·0 | 13·9 | -1·1 | -0·2 | -0·9 | -0·2 |
| 0·0040 | -19·8 | 0·0 | 11·1 | 11·1 | 11·2 | 11·9 | -0·8 | 0·0 | -0·7 | -0·1 |
| 0·0079 | -59·7 | 0·0 | 9·0 | 8·5 | 9·0 | 9·7 | -0·7 | 0·5 | -0·7 | 0·0 |
| 0·0158 | -140·5 | 0·0 | 6·9 | 5·3 | 5·7 | 7·6 | -0·7 | 1·6 | -1·9 | 1·2 |
| 0·0198 | -181·4 | 0·0 | 6·0 | 4·9 | 5·5 | 7·3 | -1·3 | 1·1 | -1·8 | 0·5 |
| 0·0395 | -390·8 | 0·0 | 4·3 | 2·9 | 3·6 | 5·4 | -1·1 | 1·4 | -1·8 | 0·7 |
| 0·0593 | -609·1 | 0·0 | 2·5 | 2·0 | 2·2 | 4·2 | -1·7 | 0·5 | -2·0 | 0·3 |
| **Injury, poisoning and certain other consequences of external causes [S00-T98] (N: 155 149, 9·5%)** | | | | | | | | | | |
| 0·0014 | 0·0 | 0·0 | 9·7 | 8·9 | 9·3 | 10·7 | -1·0 | 0·8 | -1·4 | 0·4 |
| 0·0029 | -14·5 | 0·0 | 8·4 | 7·6 | 8·0 | 9·5 | -1·1 | 0·8 | -1·5 | 0·4 |
| 0·0058 | -43·6 | 0·0 | 6·6 | 6·0 | 7·2 | 8·3 | -1·7 | 0·6 | -1·1 | -0·6 |

| | Net Benefit per 10 000 Patients | | | | | | Net Benefit Difference per 10 000 Patients | | | |
|---|---|---|---|---|---|---|---|---|---|---|
| **Risk Threshold** | **Treat All** | **Treat None** | **NEWS** | **Simplified NEWS** | **DEWS** | **XGB-EWS** | **NEWS - XGB-EWS** | **NEWS - Simplified NEWS** | **DEWS - XGB-EWS** | **NEWS - DEWS** |
| 0·0116 | -102·4 | 0·0 | 5·4 | 4·9 | 5·2 | 6·5 | -1·1 | 0·5 | -1·3 | 0·2 |
| 0·0145 | -132·1 | 0·0 | 5·0 | 4·4 | 4·7 | 6·3 | -1·3 | 0·6 | -1·6 | 0·3 |
| 0·0289 | -283·0 | 0·0 | 3·1 | 3·1 | 3·5 | 4·7 | -1·6 | 0·0 | -1·2 | -0·4 |
| 0·0434 | -438·5 | 0·0 | 2·3 | 2·4 | 2·5 | 4·1 | -1·8 | -0·1 | -1·6 | -0·2 |
| **Mental, Behavioral and Neurodevelopmental disorders [F00-F99] (N: 31 873, 1·9%)** | | | | | | | | | | |
| 0·0012 | 0·0 | 0·0 | 4·7 | 4·4 | 7·0 | 7·6 | -2·9 | 0·3 | -0·6 | -2·3 |
| 0·0023 | -11·6 | 0·0 | 4·0 | 3·1 | 6·3 | 5·9 | -1·9 | 0·9 | 0·4 | -2·3 |
| 0·0046 | -35·0 | 0·0 | 2·1 | 1·2 | 3·7 | 4·9 | -2·8 | 0·9 | -1·2 | -1·6 |
| 0·0093 | -82·0 | 0·0 | 1·2 | 1·1 | 2·7 | 4·1 | -2·9 | 0·1 | -1·4 | -1·5 |
| 0·0116 | -105·6 | 0·0 | 1·7 | 0·4 | 2·6 | 4·1 | -2·4 | 1·3 | -1·5 | -0·9 |
| 0·0232 | -225·6 | 0·0 | 1·0 | -0·4 | 1·0 | 2·0 | -1·0 | 1·4 | -1·0 | 0·0 |
| 0·0348 | -348·5 | 0·0 | 0·8 | -0·6 | 1·2 | 2·0 | -1·2 | 1·4 | -0·8 | -0·4 |
| **Neoplasms [C00-D48] (N: 66 880, 4·1%)** | | | | | | | | | | |
| 0·0027 | 0·0 | 0·0 | 17·7 | 16·3 | 14·4 | 18·6 | -0·9 | 1·4 | -4·2 | 3·3 |
| 0·0054 | -27·2 | 0·0 | 15·3 | 12·7 | 12·2 | 16·1 | -0·8 | 2·6 | -3·9 | 3·1 |
| 0·0108 | -82·1 | 0·0 | 11·0 | 9·4 | 7·9 | 12·6 | -1·6 | 1·6 | -4·7 | 3·1 |
| 0·0216 | -193·6 | 0·0 | 7·4 | 6·4 | 5·8 | 10·3 | -2·9 | 1·0 | -4·5 | 1·6 |
| 0·0271 | -250·3 | 0·0 | 6·9 | 5·7 | 3·9 | 9·3 | -2·4 | 1·2 | -5·4 | 3·0 |
| 0·0541 | -543·6 | 0·0 | 4·5 | 2·8 | 3·0 | 7·1 | -2·6 | 1·7 | -4·1 | 1·5 |
| 0·0812 | -854·1 | 0·0 | 3·6 | 2·3 | 2·3 | 5·6 | -2·0 | 1·3 | -3·3 | 1·3 |
| **Symptoms, signs and abnormal clinical and laboratory findings, not elsewhere classified [R00-R99] (N: 434 366, 26·4%)** | | | | | | | | | | |
| 0·0023 | 0·0 | 0·0 | 15·8 | 15·3 | 15·5 | 16·9 | -1·1 | 0·5 | -1·4 | 0·3 |
| 0·0047 | -23·4 | 0·0 | 13·7 | 12·6 | 13·5 | 14·6 | -0·9 | 1·1 | -1·1 | 0·2 |
| 0·0093 | -70·5 | 0·0 | 11·2 | 9·5 | 10·4 | 12·3 | -1·1 | 1·7 | -1·9 | 0·8 |
| 0·0186 | -166·1 | 0·0 | 8·0 | 7·1 | 7·9 | 10·2 | -2·2 | 0·9 | -2·3 | 0·1 |
| 0·0233 | -214·6 | 0·0 | 7·6 | 6·6 | 7·4 | 9·6 | -2·0 | 1·0 | -2·2 | 0·2 |
| 0·0466 | -464·1 | 0·0 | 5·4 | 4·4 | 4·5 | 7·4 | -2·0 | 1·0 | -2·9 | 0·9 |
| 0·0699 | -726·1 | 0·0 | 4·1 | 3·3 | 3·4 | 6·3 | -2·2 | 0·8 | -2·9 | 0·7 |
| Positive values of net benefit difference indicate the first model has higher net benefit; negative values indicate the comparator is better. Risk thresholds are chosen as multiples of the disease-specific prevalence. | | | | | | | | | | |

**Supplementary Table 8.**

Net Benefit comparison of different strategies and models across disease-chapters (ICD-10 codes) at specified risk thresholds

| | Net Benefit per 10 000 Patients | | | | | | Net Benefit Difference per 10 000 Patients | | | |
|---|---|---|---|---|---|---|---|---|---|---|
| Risk Threshold | Treat All | Treat None | NEWS | Simplified NEWS | DEWS | XGB-EWS | NEWS - XGB-EWS | NEWS - Simplified NEWS | DEWS - XGB-EWS | NEWS - DEWS |
| 0·0042 | 0·0 | 0·0 | 25·3 | 24·6 | 25·6 | 29·2 | -3·9 | 0·7 | -3·6 | -0·3 |
| 0·0083 | -42·0 | 0·0 | 20·9 | 19·5 | 21·0 | 24·8 | -3·9 | 1·4 | -3·8 | -0·1 |
| 0·0167 | -127·1 | 0·0 | 15·7 | 14·5 | 15·7 | 19·6 | -3·9 | 1·2 | -3·9 | 0·0 |
| 0·0333 | -301·7 | 0·0 | 10·1 | 8·7 | 10·3 | 14·6 | -4·5 | 1·4 | -4·3 | -0·2 |
| 0·0417 | -391·3 | 0·0 | 9·0 | 7·5 | 9·2 | 13·0 | -4·0 | 1·5 | -3·8 | -0·2 |
| 0·0833 | -863·6 | 0·0 | 4·7 | 3·4 | 4·9 | 8·3 | -3·6 | 1·3 | -3·4 | -0·2 |
| 0·1250 | -1 381·0 | 0·0 | 2·9 | 2·0 | 3·2 | 5·9 | -3·0 | 0·9 | -2·7 | -0·3 |

Positive values of net benefit difference indicate the first model has higher net benefit; negative values indicate the comparator is better. Risk thresholds are chosen as multiples of the mortality at 48 hours prevalence in the final analytic cohort (N = 2 080 765).

**Supplementary Table 9.**
Net Benefit Comparison of different strategies and models at specified risk thresholds for 48-hour mortality

| | Net Benefit per 10 000 Patients | | | | | | Net Benefit Difference per 10 000 Patients | | | |
|---|---|---|---|---|---|---|---|---|---|---|
| Risk Threshold | Treat All | Treat None | NEWS | Simplified NEWS | DEWS | XGB-EWS | NEWS - XGB-EWS | NEWS -Simplified NEWS | DEWS - XGB-EWS | NEWS - DEWS |
| 0·0060 | 0·0 | 0·0 | 36·2 | 33·9 | 36·0 | 41·5 | -5·3 | 2·3 | -5·5 | 0·2 |
| 0·0120 | -60·8 | 0·0 | 27·5 | 26·0 | 28·3 | 34·5 | -7·0 | 1·5 | -6·2 | -0·8 |
| 0·0240 | -184·7 | 0·0 | 20·0 | 18·6 | 20·6 | 26·5 | -6·5 | 1·4 | -5·9 | -0·6 |
| 0·0481 | -442·0 | 0·0 | 12·1 | 10·5 | 12·7 | 18·9 | -6·8 | 1·6 | -6·2 | -0·6 |
| 0·0601 | -575·5 | 0·0 | 10·5 | 8·8 | 11·1 | 16·5 | -6·0 | 1·7 | -5·4 | -0·6 |
| 0·1202 | -1 298·0 | 0·0 | 4·9 | 3·7 | 5·5 | 9·6 | -4·7 | 1·2 | -4·1 | -0·6 |
| 0·1803 | -2 126·4 | 0·0 | 2·8 | 1·3 | 3·3 | 6·4 | -3·6 | 1·5 | -3·1 | -0·5 |

Positive values of net benefit difference indicate the first model has higher net benefit; negative values indicate the comparator is better. Risk thresholds are chosen as multiples of the mortality at 72 hours prevalence in the final analytic cohort (N = 2 080 765).

**Supplementary Table 10.**
Net Benefit Comparison of different strategies and models at specified risk thresholds for 72-hour mortality

| | Net Benefit per 10 000 Patients | | | | | | Net Benefit Difference per 10 000 Patients | | | |
|---|---|---|---|---|---|---|---|---|---|---|
| Risk Threshold | Treat All | Treat None | NEWS | Simplified NEWS | DEWS | XGB-EWS | NEWS - XGB-EWS | NEWS - Simplified NEWS | DEWS - XGB-EWS | NEWS - DEWS |
| 0·0041 | 0·0 | 0·0 | 22·1 | 21·3 | 18·9 | 25·5 | -3·4 | 0·8 | -6·6 | 3·2 |
| 0·0082 | -41·2 | 0·0 | 18·2 | 16·9 | 16·1 | 21·1 | -2·9 | 1·3 | -5·0 | 2·1 |
| 0·0163 | -124·6 | 0·0 | 13·9 | 12·1 | 11·6 | 16·5 | -2·6 | 1·8 | -4·9 | 2·3 |
| 0·0327 | -295·6 | 0·0 | 9·3 | 7·9 | 7·4 | 12·1 | -2·8 | 1·4 | -4·7 | 1·9 |
| 0·0409 | -383·3 | 0·0 | 8·1 | 6·7 | 6·0 | 10·8 | -2·7 | 1·4 | -4·8 | 2·1 |
| 0·0817 | -845·3 | 0·0 | 4·3 | 3·1 | 3·1 | 6·8 | -2·5 | 1·2 | -3·7 | 1·2 |
| 0·1226 | -1 350·2 | 0·0 | 2·8 | 1·7 | 1·7 | 4·9 | -2·1 | 1·1 | -3·2 | 1·1 |
| Positive values of net benefit difference indicate the first model has higher net benefit; negative values indicate the comparator is better. Risk thresholds are chosen as multiples of the composite outcome (Intensive Care Unit (ICU) or Mortality at 24 hours) prevalence using the full unweighted eligible cohort (N = 2 351 828) | | | | | | | | | | |

**Supplementary Table 11.**

Net Benefit Comparison of different strategies and models at specified risk thresholds for the composite outcome of Intensive Care Unit (ICU) or mortality at 24 hours

| | Net Benefit per 10 000 Patients | | | | | | Net Benefit Difference per 10 000 Patients | | | |
|---|---|---|---|---|---|---|---|---|---|---|
| Risk Threshold | Treat All | Treat None | NEWS | Simplified NEWS | DEWS | XGB-EWS | NEWS - XGB-EWS | NEWS - Simplified NEWS | DEWS - XGB-EWS | NEWS - DEWS |
| **Cohort with Length of Stay > 60 min (N: 2 062 746, 99·1%)** | | | | | | | | | | |
| 0·0022 | 0·0 | 0·0 | 14·7 | 14·1 | 14·5 | 15·8 | -1·1 | 0·6 | -1·3 | 0·2 |
| 0·0044 | -22·3 | 0·0 | 12·4 | 11·5 | 12·3 | 13·6 | -1·2 | 0·9 | -1·3 | 0·1 |
| 0·0089 | -67·2 | 0·0 | 10·0 | 8·4 | 9·5 | 11·3 | -1·3 | 1·6 | -1·8 | 0·5 |
| 0·0178 | -158·2 | 0·0 | 6·9 | 5·9 | 6·6 | 8·9 | -2·0 | 1·0 | -2·3 | 0·3 |
| 0·0222 | -204·3 | 0·0 | 6·3 | 5·2 | 6·0 | 8·1 | -1·8 | 1·1 | -2·1 | 0·3 |
| 0·0444 | -441·4 | 0·0 | 4·1 | 2·9 | 3·6 | 5·9 | -1·8 | 1·2 | -2·3 | 0·5 |
| 0·0666 | -689·8 | 0·0 | 2·9 | 2·0 | 2·7 | 4·7 | -1·8 | 0·9 | -2·0 | 0·2 |
| Positive values of net benefit difference indicate the first model has higher net benefit; negative values indicate the comparator is better. Risk thresholds are chosen as multiples of the mortality prevalence for individuals with length of stay equal to 60 or more minutes in the final analytic cohort (N = 2 080 765). | | | | | | | | | | |

**Supplementary Table 12.**
Net Benefit comparison of different strategies and models for mortality at 24 hours for individuals with a length of stay of 60 or more minutes.

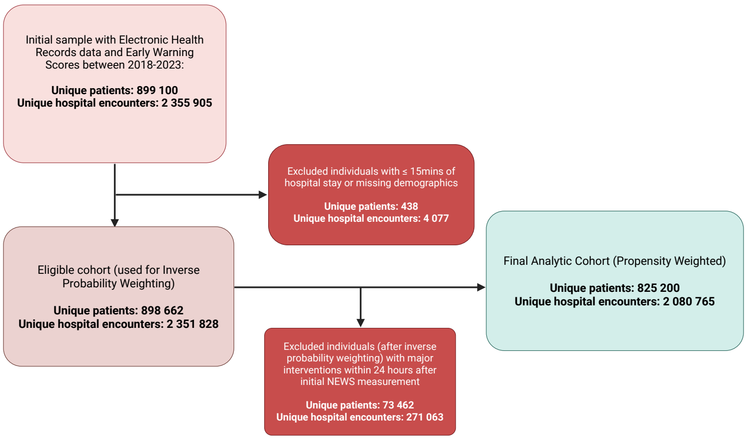

**Supplementary Figure 1.**
Sample flowchart of the current study. The flow diagram depicts the selection of the final analytic cohort from the initial dataset of 2 355 905 hospital encounters. The "Eligible cohort" represents the target population used for propensity score estimation. To address intervention bias, we employed a hypothetical strategy: patients who received major interventions within 24 hours (e.g., ICU admission, mechanical ventilation, surgery) were excluded from the outcome analysis, as their observed survival does not reflect their natural risk trajectory. The remaining "Final Analytic Cohort" was weighted using Inverse Probability Weighting (IPW) to approach the baseline risk characteristics of the full target population. The "Final Analytic Cohort" was used to develop and validate the models in the study. Created in BioRender. Katsiferis, A. (2026) https://BioRender.com/t5utde4.

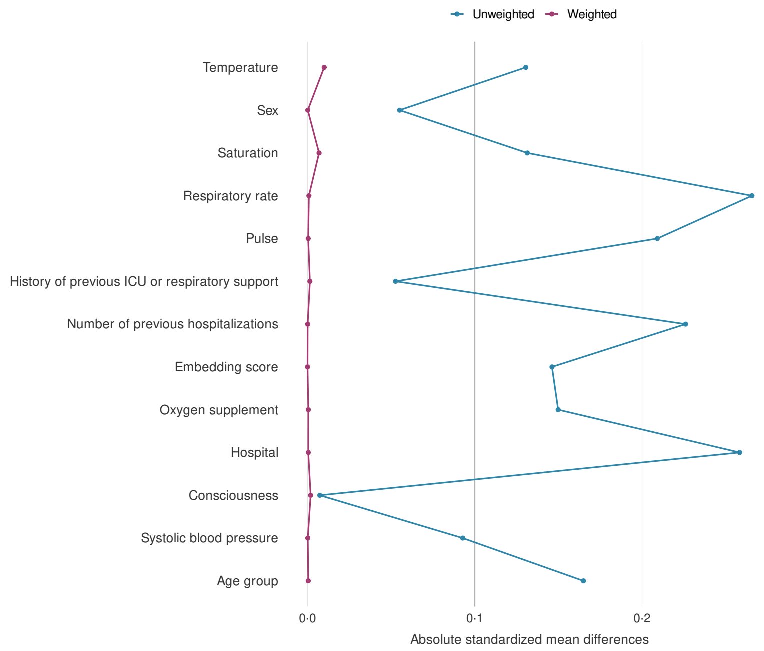

**Supplementary Figure 2.**
Covariate balance assessment before and after the weighting in the full eligible cohort. Absolute standardized mean differences between treatment groups are shown for key baseline covariates before (unweighted, blue) and after (weighted, red) weighting. Each point represents the standardized mean difference for the corresponding covariate listed on the y-axis.

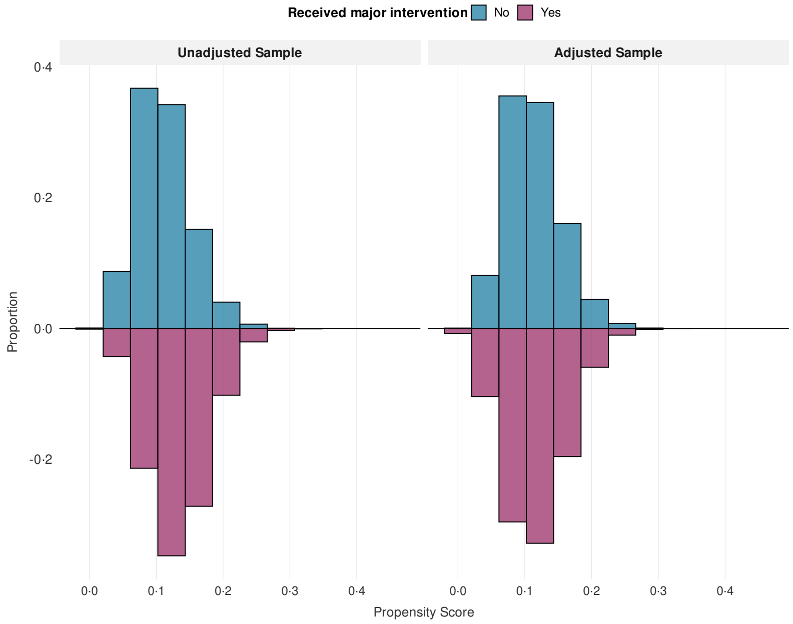

**Supplementary Figure 3.**

Distribution of propensity scores before and after inverse probability weighting in the full eligible cohort. Propensity score distributions for patients who did and did not receive major clinical interventions within 24 hours, shown before (left panel) and after (right panel) applying inverse probability weighting. Propensity scores represent the predicted probability of receiving major interventions based on baseline characteristics including age, sex, hospital, vital signs, number of previous hospitalisations, and clinical embedding scores. The mirrored histogram displays the proportion of patients (y-axis) across propensity score values (x-axis). Blue bars represent patients who did not receive major interventions; purple bars represent patients who received major interventions. Successful balancing is demonstrated by the similar distributions between groups in the weighted sample (right panel) compared to the substantial overlap reduction in the unweighted sample (left panel).

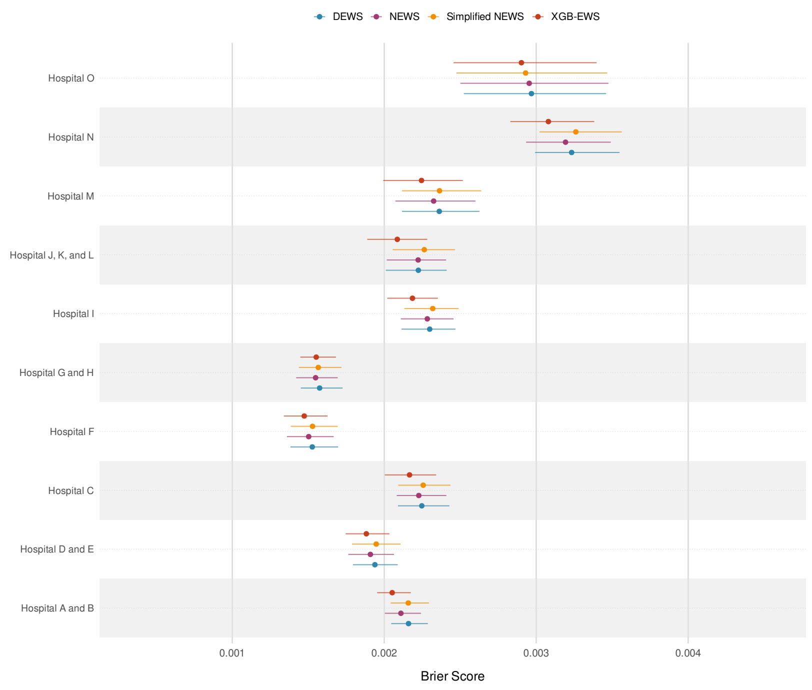

**Supplementary Figure 4.**

Forest plot of hospital-specific predictive performance of EWS algorithms. The Brier Score with 95% confidence intervals for predicting 24-hour mortality across the 10 hospital clusters is illustrated. Each point represents the Brier Score of the four prediction models within individual hospital settings. Confidence intervals were calculated using 200 bootstrap resamples with the percentile method (2·5th and 97·5th percentiles). The four models evaluated were XGB-EWS, DEWS, NEWS, and Simplified NEWS, all of which were assessed across all participating hospitals in the study's hospital network.

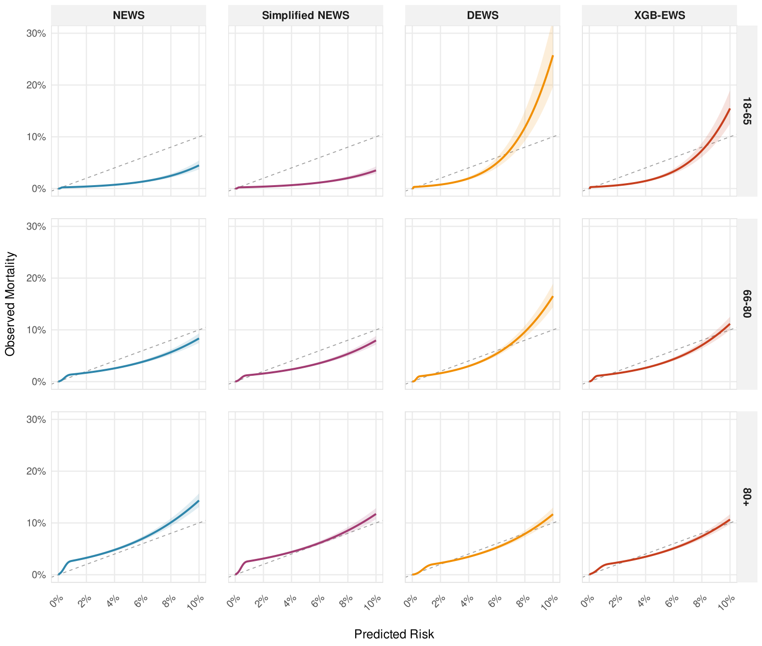

**Supplementary Figure 5.**

Model calibration by age group. The flexible calibration plots display predicted probabilities versus observed 24-hour mortality across the three age groups. The curves were estimated using logistic regression with restricted cubic splines (4 knots) to visualize calibration performance across the risk spectrum. The solid lines represent the estimated calibration curve, while the shaded areas indicate the 95% confidence intervals. The dashed diagonal line represents perfect calibration (where predicted risk equals observed risk).

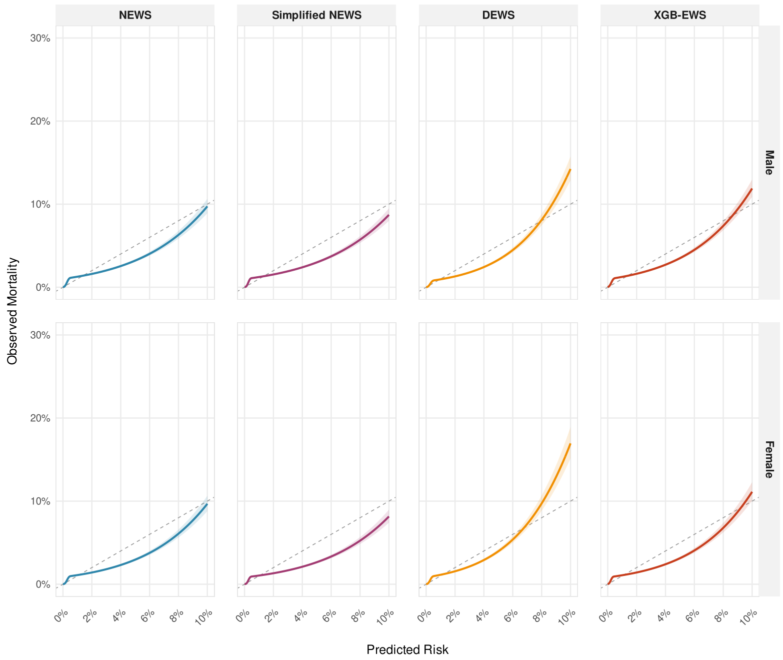

**Supplementary Figure 6.**
Model calibration across sexes. The flexible calibration plots display predicted probabilities versus observed 24-hour mortality for male and female patients. The curves were estimated using logistic regression with restricted cubic splines (4 knots) to visualize calibration performance across the risk spectrum. The solid lines represent the estimated calibration curve, while the shaded areas indicate the 95% confidence intervals. The dashed diagonal line represents perfect calibration. All estimates were calculated using inverse probability weights to correct for intervention bias and reflect the target population.

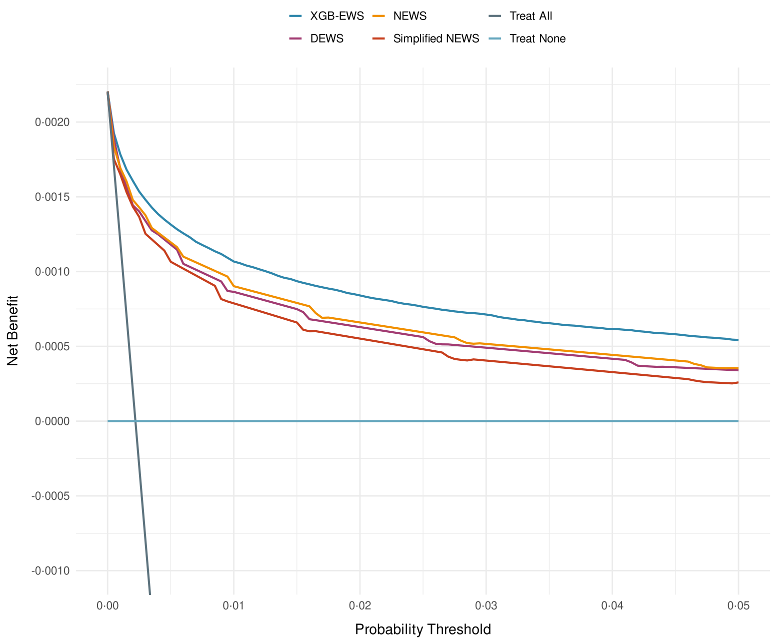

**Supplementary Figure 7.**
Decision curve analysis comparing model performance across probability thresholds. The Net Benefit is plotted against probability/risk thresholds for the four predictive models (XGB-EWS in blue, NEWS in orange, DEWS in purple, Simplified NEWS in red) compared to two reference strategies: "Treat All" (dark gray) and "Treat None" (light blue horizontal line). The Decision curve analysis evaluates the potential clinical utility of prediction models by weighing the benefits of true positive classifications against the harms of false positive classifications at different probability thresholds. Models with curves above the reference strategies indicate potential clinical utility, with higher curves representing greater Net Benefit at the corresponding threshold. The probability thresholds represent the probability cutpoints above which clinicians would initiate interventions or escalate care; for instance, a threshold of 0.01 (1%) means that patients with predicted mortality risk above 1% would receive intervention, while those below would not.

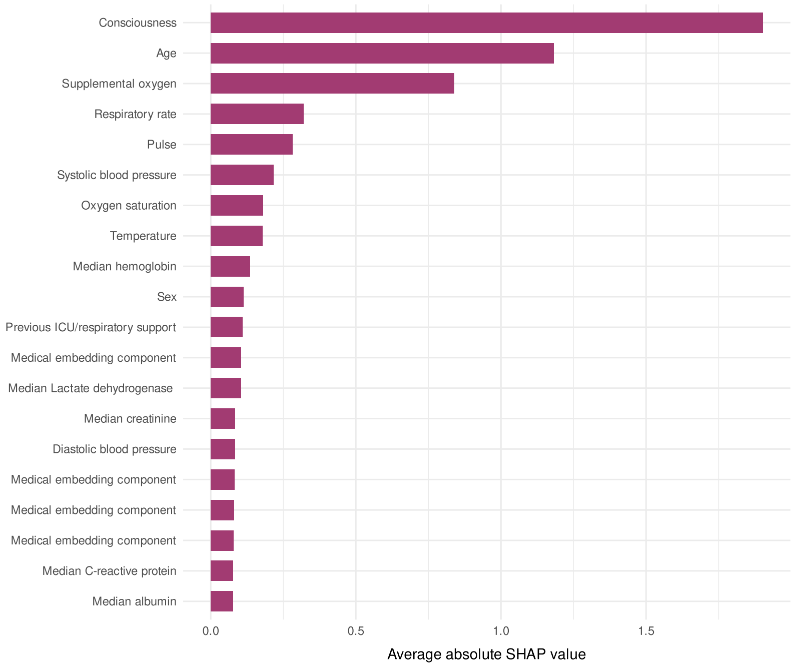

**Supplementary Figure 8.**

Top-20 predictors of the XGB-EWS model for 24-hour mortality after the time of the initial measurement of NEWS based on average absolute SHAP-values. The bars represent the average absolute SHAP-value for each predictor. The predictors are sorted in descending importance.

### Supplemental Content 3 – TRIPOD+AI Checklist

| Section/Topic | Item | Development / evaluation[1] | Checklist item | Reported on page |
|---|---|---|---|---|
| **TITLE** | | | | |
| *Title* | 1 | D;E | Identify the study as developing or evaluating the performance of a multivariable prediction model, the target population, and the outcome to be predicted | Title Page |
| **ABSTRACT** | | | | |
| *Abstract* | 2 | D;E | See TRIPOD+AI for Abstracts checklist | Abstract Page |
| **INTRODUCTION** | | | | |
| *Background* | 3a | D;E | Explain the healthcare context (including whether diagnostic or prognostic) and rationale for developing or evaluating the prediction model, including references to existing models | Page 1 (1-34) |
| | 3b | D;E | Describe the target population and the intended purpose of the prediction model in the context of the care pathway, including its intended users (e.g., healthcare professionals, patients, public) | Page 1 (36-47) |
| | 3c | D;E | Describe any known health inequalities between sociodemographic groups | Not formally assessed; subgroup analyses described on Page 4 (167-197);<br><br>Ethnicity unavailable discussed on Page 5 (239-240) |
| *Objectives* | 4 | D;E | Specify the study objectives, including whether the study describes the development or validation of a prediction model (or both) | Page 1 (36-47) |
| **METHODS** | | | | |
| *Data* | 5a | D;E | Describe the sources of data separately for the development and evaluation datasets (e.g., randomised trial, cohort, routine care or registry data), the rationale for using these data, and representativeness of the data | Page 2 (51-61) Appendix (Grouped Cross-Validation strategy section) |
| | 5b | D;E | Specify the dates of the collected participant data, including start and end of participant accrual; and, if applicable, end of follow-up | Page 2 (51-60) |
| *Participants* | 6a | D;E | Specify key elements of the study setting (e.g., primary care, secondary care, general population) including the number and location of centres | Page 2 (51-54) Appendix Grouped Cross-Validation Strategy) |
| | 6b | D;E | Describe the eligibility criteria for study participants | Page 2 (63-68) |
| | 6c | D;E | Give details of any treatments received, and how they were handled during model development or evaluation, if relevant | Page 2 (89, 93-95)<br>Page 3 (104-118)<br>Appendix (Methodological Handling of Intervention Bias section) |
| *Data preparation* | 7 | D;E | Describe any data pre-processing and quality checking, including whether this was similar across relevant sociodemographic groups | Page 2 (85-95, 97-103)<br>Appendix (Temporal Distribution & Repeat visit patterns) |
| *Outcome* | 8a | D;E | Clearly define the outcome that is being predicted and the time horizon, including how and when assessed, the rationale for choosing this outcome, and whether the method of outcome assessment is consistent across sociodemographic groups | Page 2 (70-79)<br><br>Appendix Page 1 |
| | 8b | D;E | If outcome assessment requires subjective interpretation, describe the qualifications and demographic characteristics of the outcome assessors | For post-discharge deaths, certifying physician assessment described in Appendix Page 1 and Page 5 (245-248) |
| | 8c | D;E | Report any actions to blind assessment of the outcome to be predicted | NA |
| *Predictors* | 9a | D | Describe the choice of initial predictors (e.g., literature, previous models, all available predictors) and any pre-selection of predictors before model building | Page 2 (81-95)<br><br>Page 3 (120-128)<br><br>Appendix (Description and Rationale for the Demographic Early Warning |

| | | | | |
|---|---|---|---|---|
| | | | | Score)<br><br>Appendix (Supplementary Table 2)<br><br>Appendix (Ablation Analysis & Simplified NEWS section) |
| | 9b | D;E | Clearly define all predictors, including how and when they were measured (and any actions to blind assessment of predictors for the outcome and other predictors) | Page 2 (81-95)<br><br>Appendix (Supplementary Table 2) |
| | 9c | D;E | If predictor measurement requires subjective interpretation, describe the qualifications and demographic characteristics of the predictor assessors | NA |
| *Sample size* | 10 | D;E | Explain how the study size was arrived at (separately for development and evaluation), and justify that the study size was sufficient to answer the research question. Include details of any sample size calculation | Appendix (Sample Size Considerations section)<br><br>Page 4 (149-157) for final cohort and event count |
| *Missing data* | 11 | D;E | Describe how missing data were handled. Provide reasons for omitting any data | Page 2 (94-95) Appendix (Handling of Missing Data section, Supplementary Table 1) |
| *Analytical methods* | 12a | D | Describe how the data were used (e.g., for development and evaluation of model performance) in the analysis, including whether the data were partitioned, considering any sample size requirements | Page 3 (130-139) Appendix (Grouped Cross-Validation Strategy) Appendix (Sample Size Considerations) |
| | 12b | D | Depending on the type of model, describe how predictors were handled in the analyses (functional form, rescaling, transformation, or any standardisation). | Page 2 (97-103)<br><br>Page 3 (120-128)<br><br>Appendix (Ablation Analysis & Simplified NEWS section, Description and Rationale for the Demographic Early Warning Score section) |
| | 12c | D | Specify the type of model, rationale[2], all model-building steps, including any hyperparameter tuning, and method for internal validation | Page 3 (120-139) Appendix (Grouped Cross-Validation Strategy, Supplementary Table 2, Supplementary Table 3) |
| | 12d | D;E | Describe if and how any heterogeneity in estimates of model parameter values and model performance was handled and quantified across clusters (e.g., hospitals, countries). See TRIPOD-Cluster for additional considerations[3] | Page 3 (130-139) Appendix (Grouped Cross-Validation Strategy) |
| | 12e | D;E | Specify all measures and plots used (and their rationale) to evaluate model performance (e.g., discrimination, calibration, clinical utility) and, if relevant, to compare multiple models | Page 3 (130-139) |
| | 12f | E | Describe any model updating (e.g., recalibration) arising from the model evaluation, either overall or for particular sociodemographic groups or settings | NA |
| | 12g | E | For model evaluation, describe how the model predictions were calculated (e.g., formula, code, object, application programming interface) | Page 6 (282-283)<br><br>Appendix model specification sections |
| *Class imbalance* | 13 | D;E | If class imbalance methods were used, state why and how this was done, and any subsequent methods to recalibrate the model or the model predictions | NA |
| *Fairness* | 14 | D;E | Describe any approaches that were used to address model fairness and their rationale | No formal fairness mitigation;<br><br>Subgroup analyses by age, |

| | | | | |
|---|---|---|---|---|
| | | | | sex, hospital, specialty, diagnosis, and NEWS risk strata reported on Page 3 (137-138)<br><br>Page 4 (167-197)<br><br>Figure 4<br><br>Appendix tables/figures<br><br>Ethnicity limitation discussed on Page 5 (239-240) |
| *Model output* | 15 | D | Specify the output of the prediction model (e.g., probabilities, classification). Provide details and rationale for any classification and how the thresholds were identified | Page 3 (104-108, 130-139)<br><br>Appendix (Supplementary Table 4) |

[1] D=items relevant only to the development of a prediction model; E=items relating solely to the evaluation of a prediction model; D;E=items applicable to both the development and evaluation of a prediction model

[2] Separately for all model building approaches.

[3] TRIPOD-Cluster is a checklist of reporting recommendations for studies developing or validating models that explicitly account for clustering or explore heterogeneity in model performance (eg, at different hospitals or centres). Debray et al, BMJ 2023; 380: e071018 [DOI: 10.1136/bmj-2022-071018]

| | | | | |
|---|---|---|---|---|
| *Training versus evaluation* | 16 | D;E | Identify any differences between the development and evaluation data in healthcare setting, eligibility criteria, outcome, and predictors | All models evaluated using grouped ten-fold hospital-clustered cross-validation; No separate external evaluation dataset<br><br>Page 3 (130-139)<br><br>Appendix (Grouped Cross-Validation Strategy) |
| *Ethical approval* | 17 | D;E | Name the institutional research board or ethics committee that approved the study and describe the participant-informed consent or the ethics committee waiver of informed consent | Page 3 (141-143) |
| **OPEN SCIENCE** | | | | |
| *Funding* | 18a | D;E | Give the source of funding and the role of the funders for the present study | Page 3 (144-146)<br><br>Page 6 (285-294) |
| *Conflicts of interest* | 18b | D;E | Declare any conflicts of interest and financial disclosures for all authors | Page 6 (270-276) |
| *Protocol* | 18c | D;E | Indicate where the study protocol can be accessed or state that a protocol was not prepared | Not Prepared |
| *Registration* | 18d | D;E | Provide registration information for the study, including register name and registration number, or state that the study was not registered | Not Registered |
| *Data sharing* | 18e | D;E | Provide details of the availability of the study data | Page 6 (277-283) |
| *Code sharing* | 18f | D;E | Provide details of the availability of the analytical code[4] | Page 6 (282-283)<br><br>Appendix (Model specification sections)<br><br>https://github.com/MLGlobalHealth/Early_Warning_Scores |
| **PATIENT & PUBLIC INVOLVEMENT** | | | | |
| *Patient & Public Involvement* | 19 | D;E | Provide details of any patient and public involvement during the design, conduct, reporting, interpretation, or dissemination of the study or state no involvement. | Not Reported |
| **RESULTS** | | | | |

| | | | | |
|---|---|---|---|---|
| *Participants* | 20a | D;E | Describe the flow of participants through the study, including the number of participants with and without the outcome and, if applicable, a summary of the follow-up time. A diagram may be helpful. | Page 4 (149-157) Appendix (Supplementary Figure 1) |
| | 20b | D;E | Report the characteristics overall and, where applicable, for each data source or setting, including the key dates, key predictors (including demographics), treatments received, sample size, number of outcome events, follow-up time, and amount of missing data. A table may be helpful. Report any differences across key demographic groups. | Page 4 (149-160) Table 1 Appendix (Supplementary Table 1) |
| | 20c | E | For model evaluation, show a comparison with the development data of the distribution of important predictors (demographics, predictors, and outcome). | Table 1 |
| *Model development* | 21 | D;E | Specify the number of participants and outcome events in each analysis (e.g., for model development, hyperparameter tuning, model evaluation) | Page 4 (149-157) |
| *Model specification* | 22 | D | Provide details of the full prediction model (e.g., formula, code, object, application programming interface) to allow predictions in new individuals and to enable third-party evaluation and implementation, including any restrictions to access or re-use (e.g., freely available, proprietary)[5] | Page 3 (120-128)<br>Page 6 (282-283)<br>Appendix (Supplementary Table 2-3) |
| *Model performance* | 23a | D;E | Report model performance estimates with confidence intervals, including for any key subgroups (e.g., sociodemographic). Consider plots to aid presentation. | Page 4 (162-197)<br>Figures 2-4<br>Appendix Supplementary Figures 4-7 and Supplementary Tables 5-12 |
| | 23b | D;E | If examined, report results of any heterogeneity in model performance across clusters. See TRIPOD Cluster for additional details[3]. | Figure 2, Appendix (Supplementary Figure 4) |
| *Model updating* | 24 | E | Report the results from any model updating, including the updated model and subsequent performance | NA |
| **DISCUSSION** | | | | |
| *Interpretation* | 25 | D;E | Give an overall interpretation of the main results, including issues of fairness in the context of the objectives and previous studies | Page 5 (200-233)<br>Page 6 (260-263) |
| *Limitations* | 26 | D;E | Discuss any limitations of the study (such as a non-representative sample, sample size, overfitting, missing data) and their effects on any biases, statistical uncertainty, and generalizability | Page 5 (234-250) |
| *Usability of the model in the context of current care* | 27a | D | Describe how poor quality or unavailable input data (e.g., predictor values) should be assessed and handled when implementing the prediction model | Appendix, Handling of Missing Data Section; Supplementary Table 1;<br>Page 2 (90-95) |
| | 27b | D | Specify whether users will be required to interact in the handling of the input data or use of the model, and what level of expertise is required of users | Page 5 (231-232)<br>Page 5 (235-237) |
| | 27c | D;E | Discuss any next steps for future research, with a specific view to applicability and generalizability of the model | Page 5 (226-228)<br>Page 6 (260-263) |

From: Collins GS, Moons KGM, Dhiman P, et al. *BMJ* 2024;385:e078378. doi:10.1136/bmj-2023-078378

[4] This relates to the analysis code, for example, any data cleaning, feature engineering, model building, evaluation.

[5] This relates to the code to implement the model to get estimates of risk for a new individual.
